# The Individual and Social Determinants of COVID-19 Diagnosis in Ontario, Canada: A Population-Wide Study

**DOI:** 10.1101/2020.11.09.20223792

**Authors:** Maria E. Sundaram, Andrew Calzavara, Sharmistha Mishra, Rafal Kustra, Adrienne K. Chan, Mackenzie A. Hamilton, Mohamed Djebli, Laura C. Rosella, Tristan Watson, Hong Chen, Branson Chen, Stefan D. Baral, Jeffrey C. Kwong

## Abstract

**Background:** Optimizing the public health response to reduce coronavirus disease 2019 (COVID-19) burden necessitates characterizing population-level heterogeneity of COVID-19 risks. However, heterogeneity in severe acute respiratory syndrome coronavirus 2 (SARS-CoV-2) testing may introduce biased estimates depending on analytic design.

**Methods:** We explored the potential for collider bias and characterized individual, environmental, and social determinants of testing and diagnosis using cross-sectional analyses among 14.7 million community-dwelling people in Ontario, Canada. Among those diagnosed, we used separate analytic designs to compare predictors of: 1) individuals testing positive versus negative; 2) symptomatic individuals only testing positive versus testing negative; and 3) individuals testing positive versus individuals not testing positive (i.e., testing negative or not being tested). Analyses included tests conducted between March 1 and June 20, 2020.

**Results:** Of a total of 14,695,579 individuals, 758,691 were tested for SARS-CoV-2, of whom 25,030 (3.3%) tested positive. The further the odds of testing from the null, the more variability observed in the odds of diagnosis across analytic design, particularly among individual factors. There was less variability in testing by social determinants across analytic designs. Residing in areas with highest household density (adjusted odds ratio [aOR]: 1.86; 95%CI: 1.75-1.98), highest proportion of essential workers (aOR: 1.58; 95%CI: 1.48-1.69), lowest educational attainment (aOR: 1.33; 95%CI: 1.26-1.41), and highest proportion of recent immigrants (aOR: 1.10; 95%CI: 1.05-1.15) were consistently related to increased odds of SARS-CoV-2 diagnosis regardless of analytic design.

**Interpretation:** Where testing is limited, risk factors may be better estimated using population comparators rather than test-negative comparators. Optimizing COVID-19 responses necessitates investment and sufficient coverage of structural interventions tailored to heterogeneity in social determinants of risk, including household crowding, occupation, and structural racism.

## Introduction

The spread of severe acute respiratory syndrome coronavirus 2 (SARS-CoV-2), the virus causing coronavirus disease 2019 (COVID-19), has resulted in a pandemic with heterogeneity in exposure and transmission risks.^1-4^

Heterogeneity in social determinants of COVID-19 may exist at the individual and network (for example, by housing density^5-7^) levels. In addition, social determinants of health, including barriers to healthcare, occupation, structural racism, and xenophobia have been implicated in COVID-19 risk.^8,9^ Environmental determinants such as ambient air pollution may also play a role, as existing evidence indicates that higher ambient air pollution increases risk for infection with other respiratory viruses^10,11^ and severe COVID-19.^12,13^ It could also play a role in COVID-19 risk by operating within the context of low-quality housing and structural racism.^14,15^

Using observational data to identify determinants of COVID-19 relies on SARS-CoV-2 testing, a service which is not equally distributed.^16^ Differential testing introduces the potential for selection biases,^17,18^ including collider bias.^18^ Collider bias may be introduced into epidemiological studies of COVID-19 risk factors if the factors under investigation are related to infection and to the likelihood of being tested.^18-20^ For example, data suggest individuals with diabetes are more likely to develop severe COVID-19 if infected.^21,22^ Thus, if infected, individuals with diabetes may be more likely to be tested, and consequently diabetes may appear to be associated with a diagnosis of COVID-19 in studies of individuals tested for SARS-CoV-2, even if diabetes is not a mechanistic risk factor for infection.^18^ The opposite may occur with underlying respiratory diseases (e.g., asthma) that have symptoms similar to those caused by SARS-CoV-2, leading to the appearance of potentially ‘protective’ associations with COVID-19.^23^

Our objectives were to: 1) explore the potential for collider bias in large-scale studies of COVID-19 determinants; and 2) examine individual, environmental, and social determinants associated with testing and diagnosis among 14.7 million individuals in Ontario, Canada.^18^

## Methods

### Study population, setting, and design

We conducted an observational study using population-based laboratory and health administrative databases from Ontario. Ontario’s single-payer health system provides universal access to hospital and physician services^24^ and laboratory testing.^25^ We used data from individuals tested between March 1 and June 20, 2020 to identify determinants associated with testing, and then used different analytic designs to identify determinants associated with diagnosis.

### Data sources, linkages, and inclusion criteria

We identified testing status using data from the Ontario Laboratories Information System (OLIS), and linked this information to relevant health-related datasets containing demographic, healthcare use, and area-level information. These datasets were linked using unique encoded identifiers and analyzed at ICES.^26^

OLIS captured approximately 88% of all laboratory-identified COVID-19 reported by the province during the study period (calculated by cases identified in OLIS divided by the number of cases reported by Ontario’s COVID-19 dashboard in the same time frame). OLIS records included specimen collection date, results, and a text field for symptoms completed by healthcare providers at the time of sampling. We obtained individual- and area-level demographic and environmental information from the Registered Persons Database; the Canadian Institute for Health Information’s Discharge Abstract Database, Same Day Surgery Database, and National Ambulatory Care Reporting System; the Ontario Health Insurance Program; the Ontario Mental Health Reporting System; the Ontario Population Health and Environment Cohort; and the 2016 Canadian Census^27^ (**Supplemental Table 1**).

For individuals with more than one test in OLIS, we used the first positive or indeterminate test, or the first negative test if all tests during the study period were negative. Never-tested individuals during the study period were included if they were not recorded as deceased before, or born after, March 1, 2020. To assess determinants of testing and diagnosis, we included individuals who were tested for SARS-CoV-2 by polymerase chain reaction tests and were not residing in a long-term care facility as of March 1, 2020.

### Selection and definition of potential determinants of COVID-19

Individual-level determinants included sex, age group, underlying health conditions, and prior healthcare use. Underlying health conditions were selected as: 1) health conditions identified in the literature as associated with COVID-19 severity^2,28-31^ or associated with symptoms similar to those caused by COVID-19, because severity and symptoms may lead to differential testing and thus, collider bias;^32-37^ or 2) health conditions such as dementia that increase the need for personal care support, thereby reflecting an intersection with occupational risks among essential care providers.^38,39^

Healthcare use was hypothesized to increase access to testing, and/or signal a marker for comorbidities, and was measured by number of hospital admissions in the past 3 years, number of outpatient physician visits in the past year, and influenza vaccination in the 2019-2020 season. We also included ACG^®^ System^40^ Aggregated Diagnostic Groups (ADGs)^41^ as a composite measure of comorbidities.

Environmental determinants included fine particulate matter (PM_2.5_) using satellite-derived estimates^42^ and land-use regression model for NO_2_,^43^ at the postal-code level.

Social determinants were conceptualized as area-based variables that may signal contact rates in communities [household density, apartment building density, uncoupled (e.g., not married) status];^44,45^ contact rates at work (“essential workers”);^17,46^ socio-economic barriers to healthcare access and/or housing (household income and educational attainment);^47,48^ and factors related to race/ethnicity (visible minority status and recent immigration).^8,9^ These variables were derived from the 2016 Canada Census at the level of dissemination areas (DA), the smallest geographic unit that Census data are disseminated.^49^ DAs were ranked at the city level (for median per-person income equivalent) or at the province level (for all other social determinants), and then categorized into quintiles. For apartment building density and recent immigration status, the high frequency of zeros permitted the creation of only three categories (i.e., comparing the fourth and the fifth quintiles with the lower three quintiles).

### Statistical analysis

The testing outcome was defined as receipt of at least one SARS-CoV-2 test during the study period. The comparator group comprised Ontario residents who had no record of testing during the study period. Determinants of testing were examined in unadjusted, age/sex-adjusted, and fully-adjusted logistic regression models that included all determinants. The fully-adjusted model also included a fixed-effect covariate for public health region. Public health regions are geographic areas in which public health measures were differentially applied^50^ and along which there may be variability in measured and unmeasured social determinants.^51^

To address objective 1, we compared the odds of diagnosis via unadjusted, age/sex-adjusted, and fully-adjusted logistic regression models (including all determinants and public health regions) using three study designs. The “pseudo-test-negative design” compared individuals who tested positive to individuals who tested negative; the “true test-negative design” was restricted to tested individuals with symptomatic illness;^52^ and the “case-control” design compared all individuals who tested positive to all individuals who did *not* test positive (i.e., individuals who tested negative or were not tested).

To address objective 2, we focused on results from fully-adjusted logistic regression models from the pseudo-test-negative and case-control designs. We interpreted each set of determinants as independent analyses based on directed acyclic graphs (**Supplemental Figure 1**), with the case-control design having the least potential for collider bias based on our proposed causal diagrams.

Statistical analyses were conducted using SAS v.9.4 (Cary, NC). To assess for collinearity, we examined tolerances and variance inflation factors.

### Ethical review

The use of data in this project was authorized under section 45 of Ontario’s Personal Health Information Protection Act, which does not require review by a Research Ethics Board.

## Results

Of 758,691 individuals tested during the study period, 25,030 (3.3%) were diagnosed with COVID-19 (**Figure 1**). Only 11.8% of those tested had a symptom recorded by the provider, 13.6% were considered asymptomatic, and 74.6% were missing symptom information. Descriptive characteristics of the study population are reported in **Table 1** and **Supplemental Table 2**.

**Figure 1.**
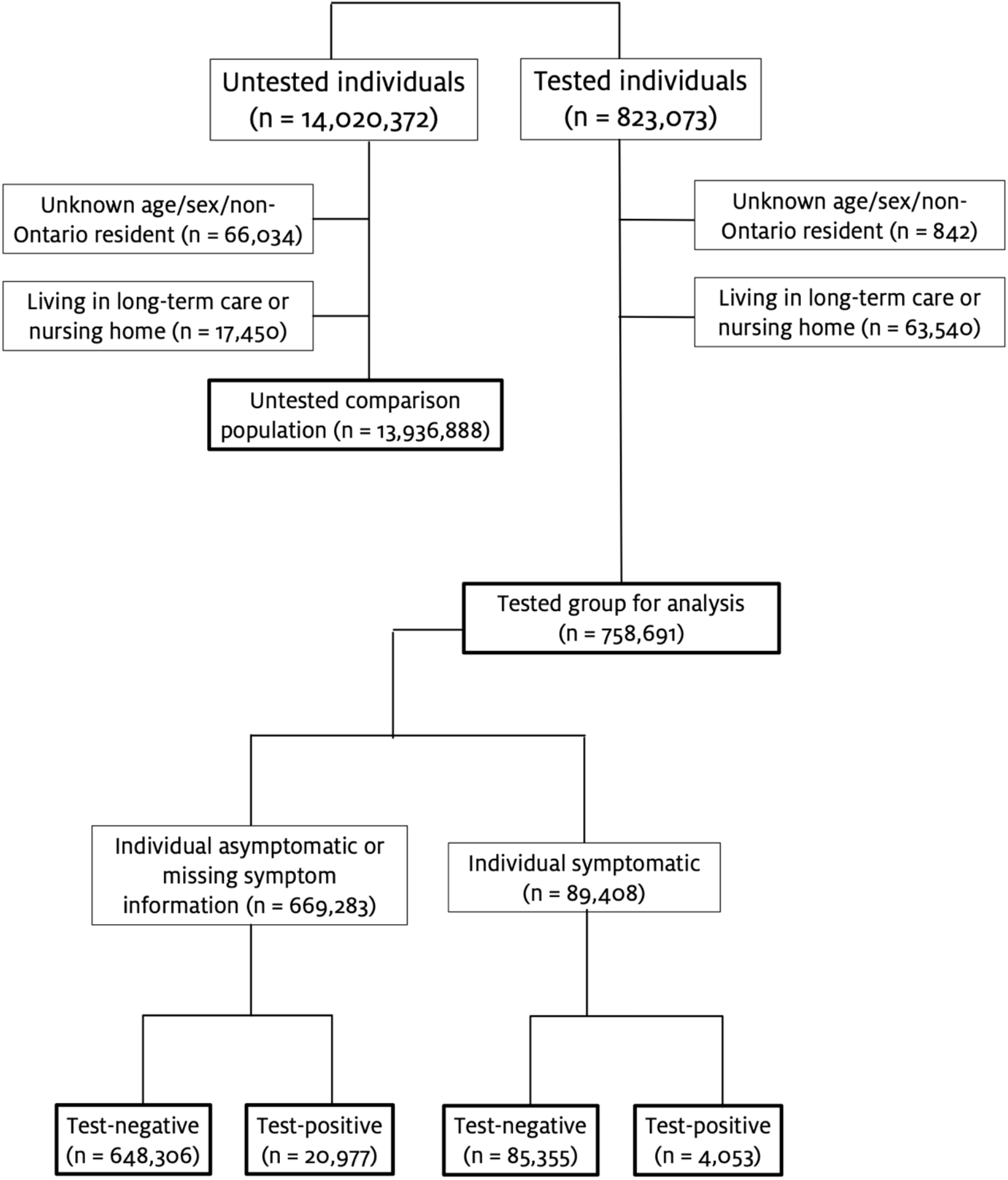
Diagram of inclusion and exclusion criteria and resulting analytic datasets. Individuals were excluded if they had a listed age >105 years, had a listed postal code outside of Ontario, or had no record of contact with the healthcare system in the past 8 years (since March 2012).

**Table 1.**
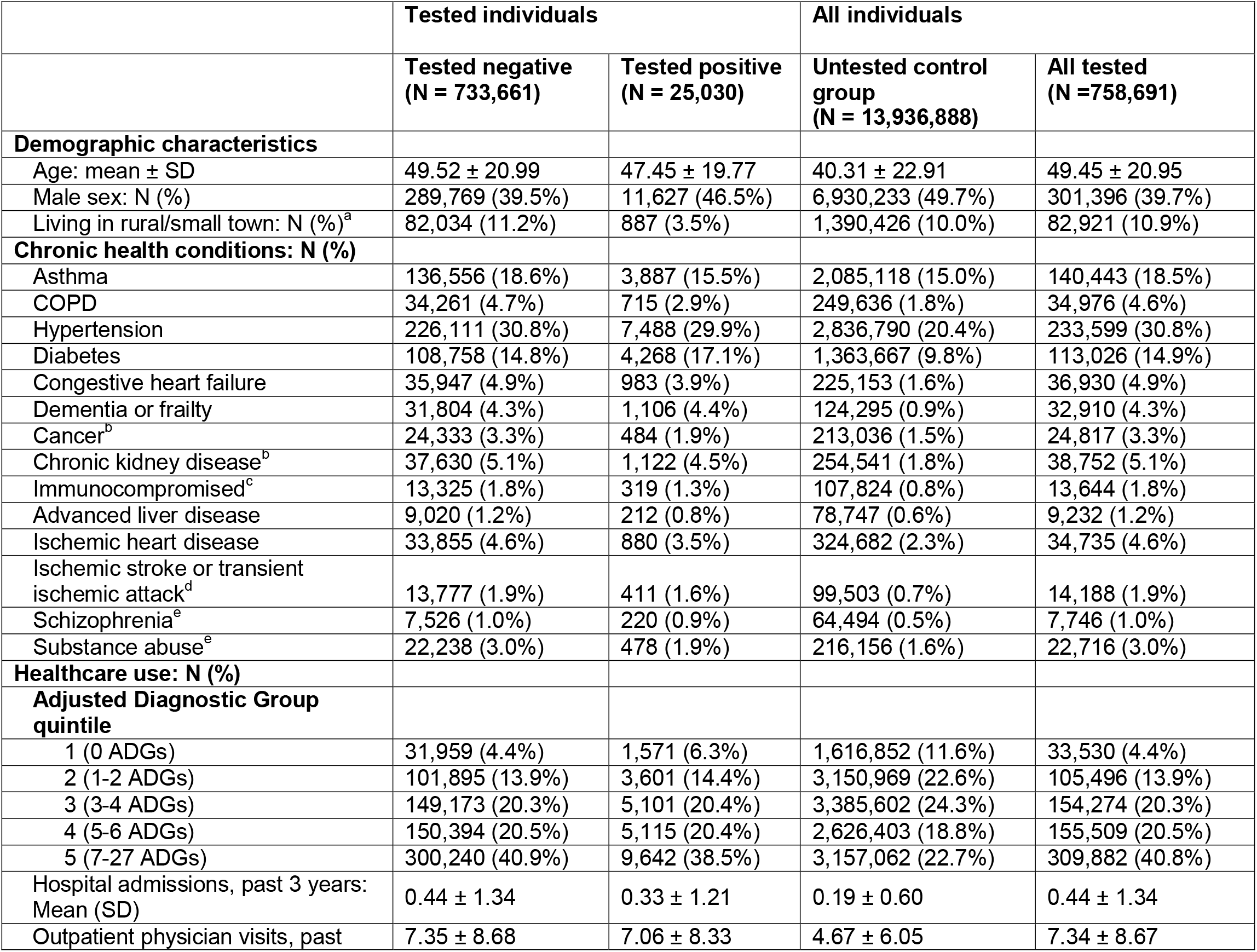
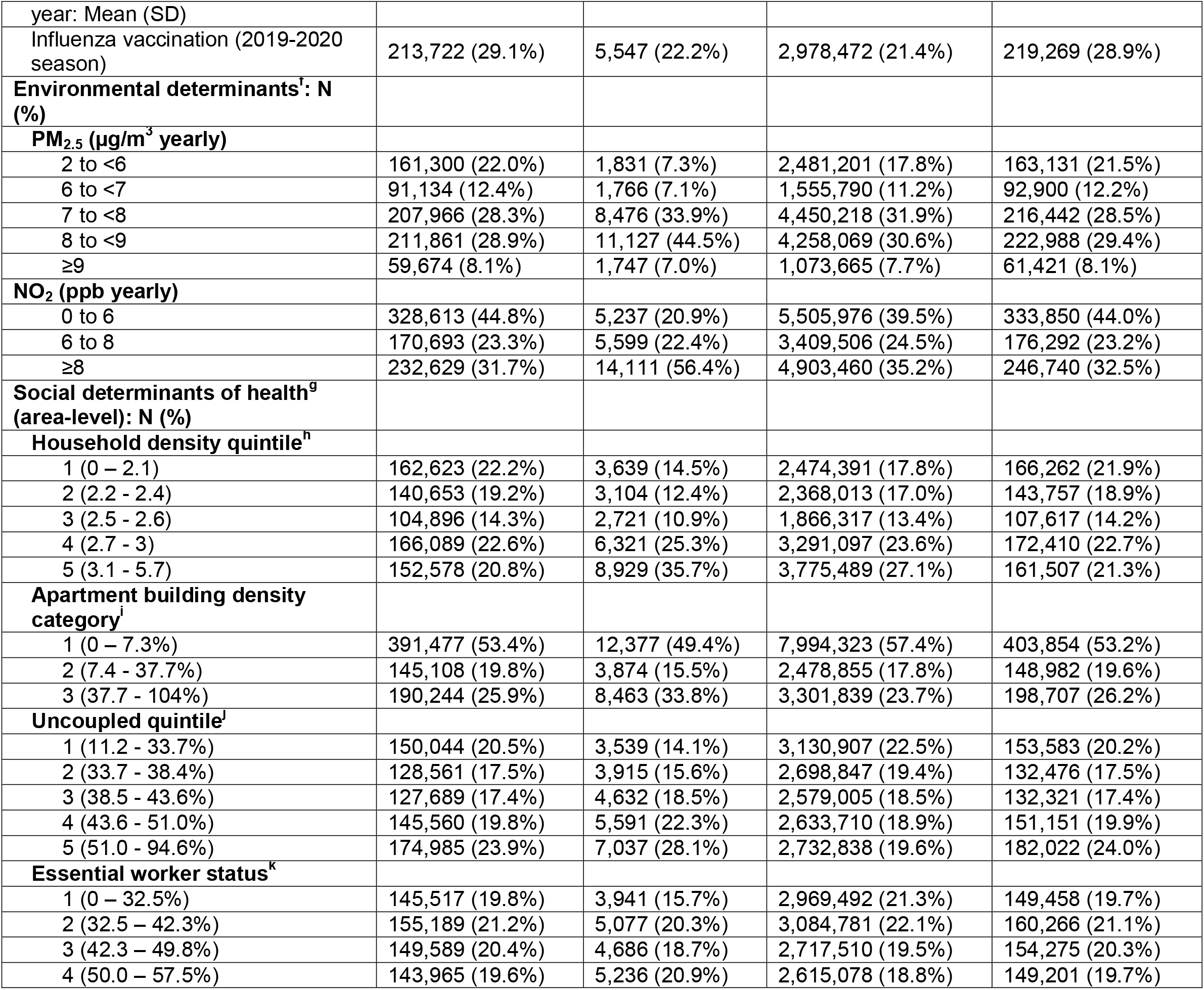
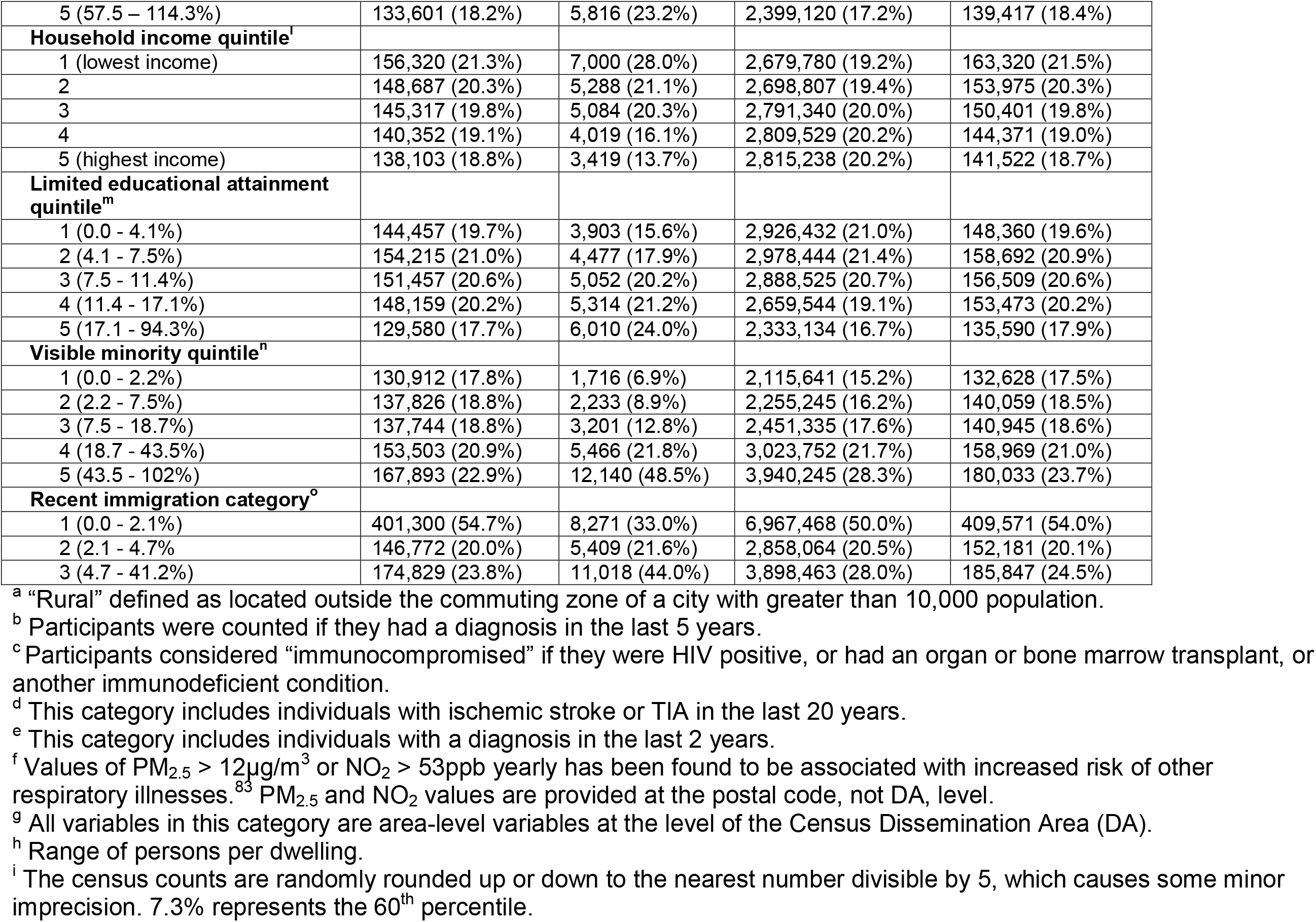
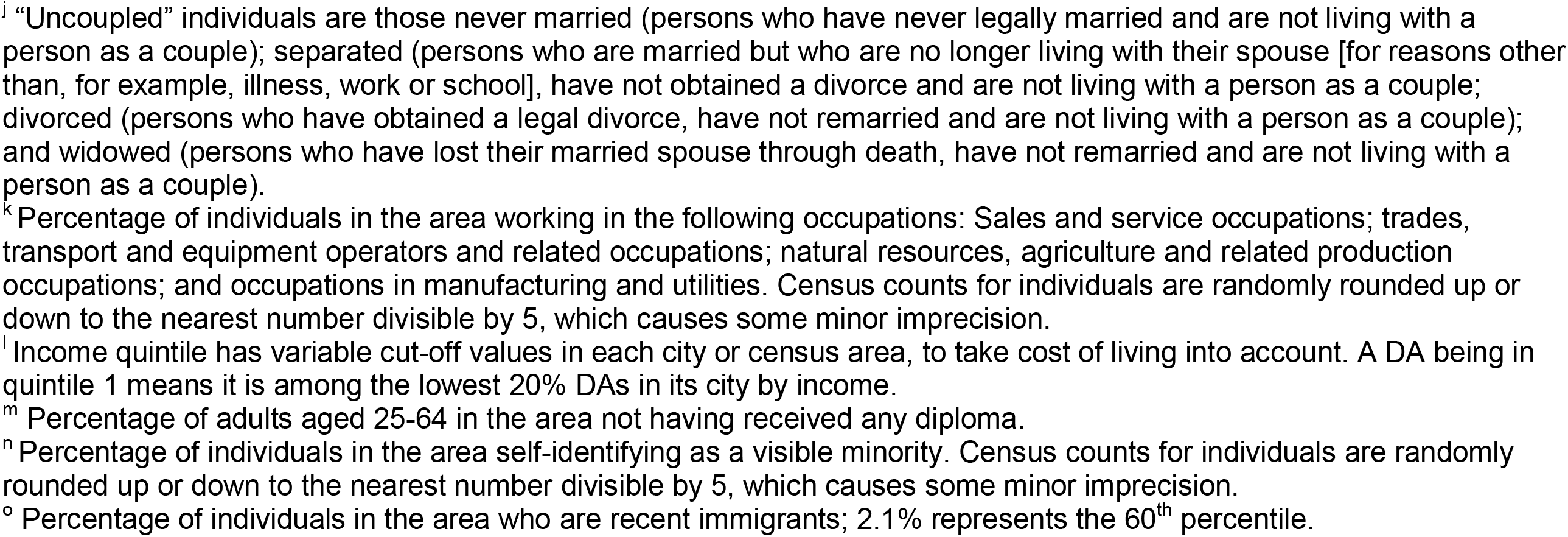
Characteristics of study population tested and not tested for SARS-CoV-2 in Ontario (March 1, 2020 to June 20, 2020).

### Determinants of testing

In fully-adjusted analyses, odds of testing increased with age (**Table 2, Supplemental Table 3**). Males had lower odds of testing than females. Nearly every underlying health condition and most measures of prior healthcare use were associated with increased odds of testing. In contrast, higher ambient air pollution was associated with reduced odds of testing. There was little variability in the odds of testing by most area-based social determinants of health. However, areas with higher visible minority populations had lower odds of testing, whereas areas with higher household income and greater percentages of uncoupled individuals had higher odds of testing. Effect measures for most social determinants of health appeared to be progressively attenuated from unadjusted, to age/sex-adjusted, to fully-adjusted regression models (**Figure 2A, Supplemental Table 3**).

**Table 2.**
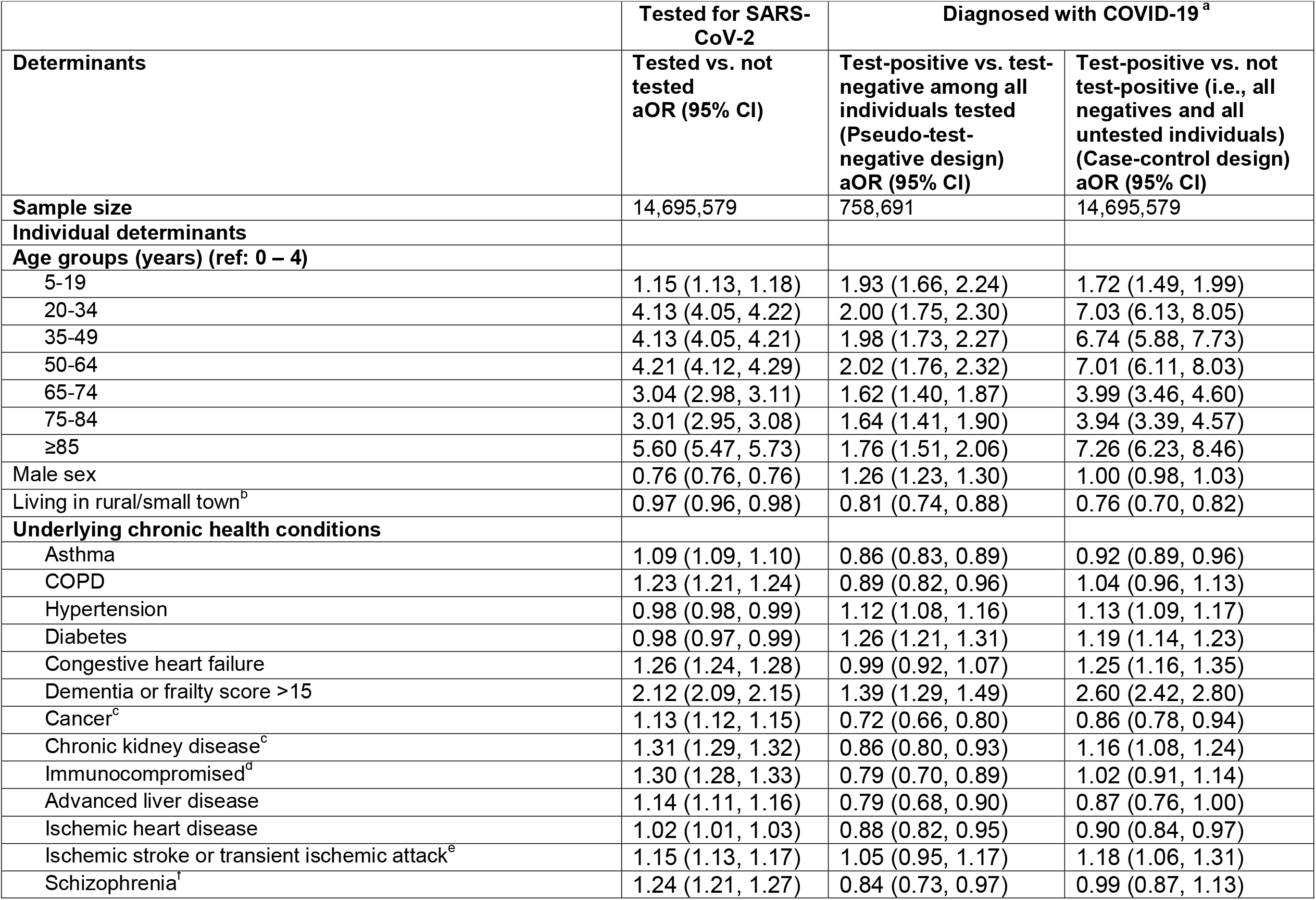
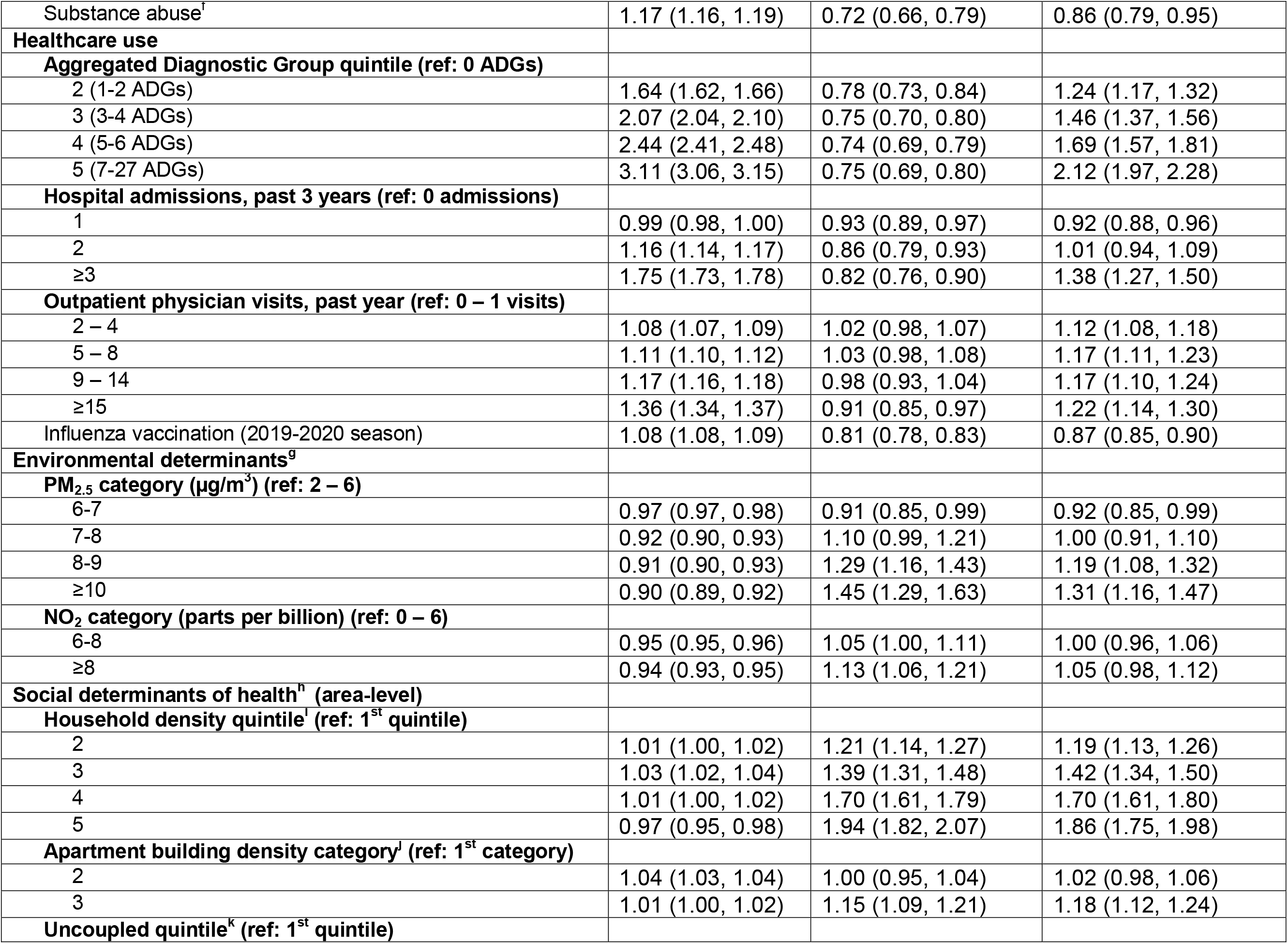
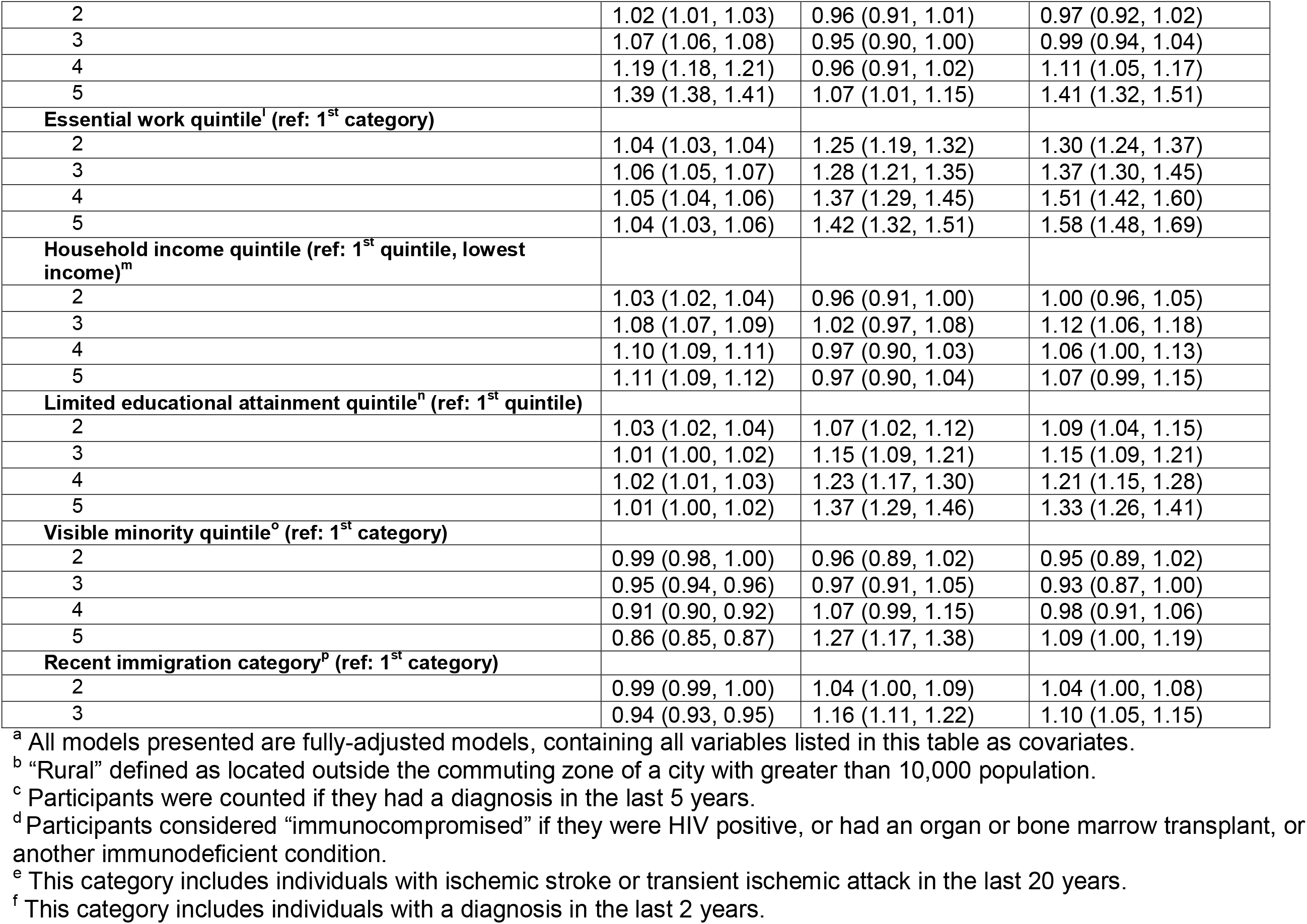
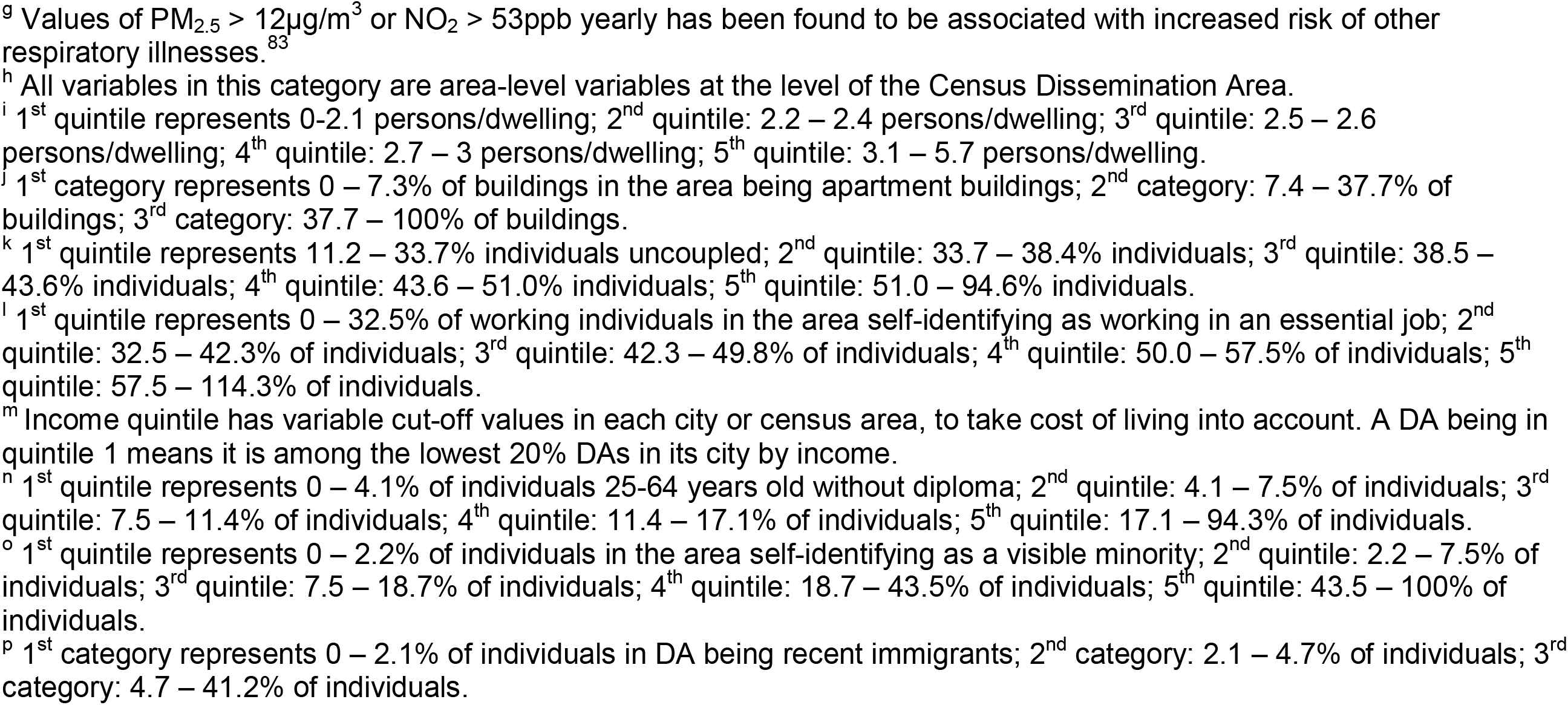
Odds of ever being tested for SARS-CoV-2 and of COVID-19 diagnosis in fully-adjusted analyses, using two analytic designs, in Ontario, Canada between March 1 and June 20, 2020.

**Figure 2.**
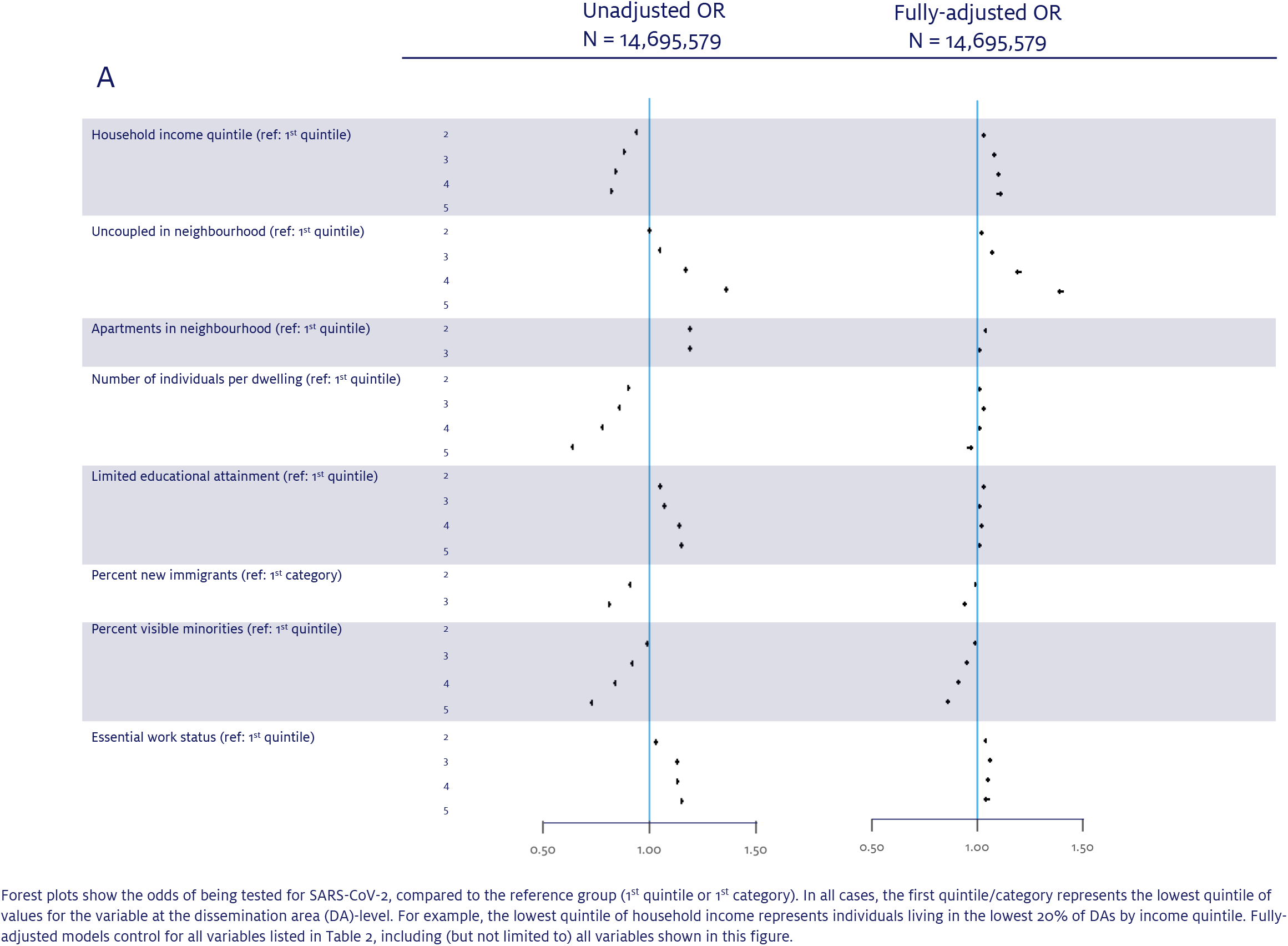
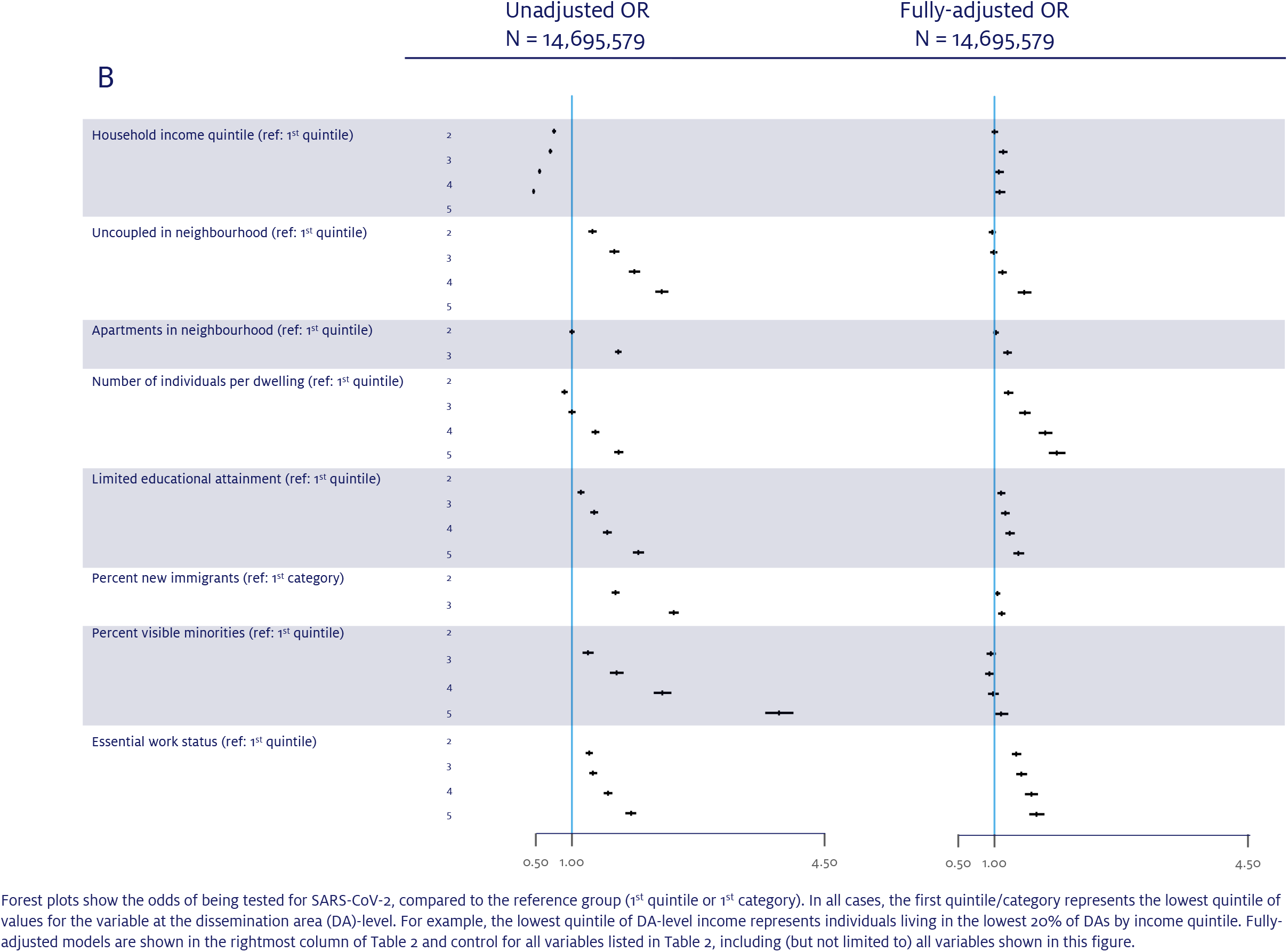
Unadjusted and fully-adjusted association between social determinants and SARS-CoV-2 testing (panel A) and COVID-19 diagnosis (panel B), using the case-control design, in Ontario (March 1 to June 20, 2020).

### Variability in determinants of diagnosis across analytic designs

Comparison of analytic designs revealed an important influence on individual-level determinant effect measures, and less of an influence on social determinants (**Table 2, Supplemental Tables 4, 5, and 6)**. Variables associated with testing tended towards a variable relationship with diagnosis, depending on the analytic design. For example, the odds of testing for adults aged ≥85 years compared to those aged <5 years deviated considerably from the null (adjusted odds ratio [aOR]=5.60; 95%CI, 5.47-5.73), and the odds of diagnosis differed between the pseudo-test-negative design (aOR=1.76; 95%CI, 1.51-2.06) and the case-control design (aOR=7.26; 95%CI, 6.23-8.46) (**Table 2**). Some health conditions associated with higher odds of testing, such as chronic respiratory conditions and indicators of prior healthcare use, appeared protective against diagnosis using the pseudo-test-negative design, but reverted to the null or showed increased odds of diagnosis using the case-control design. Results from the true test-negative design were largely similar to results from the pseudo-test-negative design with wider confidence intervals, with the exceptions that odds of diagnosis were higher for older ages using the true test-negative design compared to the pseudo-test-negative design, and lower for higher quintiles of essential workers in the true test-negative design compared to the pseudo-test-negative design (**Supplemental Tables 4 and 5**).

### Determinants of diagnosis using the case-control design

Using the case-control design, older age, certain comorbidities (hypertension, diabetes, congestive heart failure, dementia, chronic kidney disease, and ischemic stroke/transient ischemic attack), and increased prior healthcare use were associated with increased odds of diagnosis. In contrast, other comorbidities (asthma, cancer, ischemic heart disease, substance abuse) and receipt of influenza vaccine in the 2019-2020 season were associated with reduced odds of diagnosis (**Table 2, Supplemental Table 6**).

The two highest categories of PM_2.5_ exposure were associated with increased odds of diagnosis, whereas no categories of exposure to NO_2_ were associated with increased odds of diagnosis.

Higher household density, increased apartment building density, greater percentages of uncoupled individuals, and greater percentages of essential workers were associated with greater risk of diagnosis. Lower educational attainment was related to an increased risk of diagnosis, but there was no statistically consistent relationship with household income. Finally, being in the highest quintile of neighbourhoods with visible minorities, and greater percentages of recent immigrants were associated with greater odds of diagnosis. Associations were attenuated after adjustment for all social determinants except for household density and essential work (**Figure 2B, Supplemental Table 6**).

In collinearity diagnostics, all tolerances were below 1 and all variance inflation factors were below 5 (**Supplemental Table 7**).

## Discussion

We identified variability in associations by analytic design, likely due to collider bias, across individual determinants of diagnosis. Using results from the case-control analysis, we identified particular individual, environmental, and social determinants of health as key determinants for diagnosis.

We found a high potential for erroneously identifying a “risk/protective factor” with individual determinants such as underlying health conditions, sex, and age. Our findings identified a pattern where underlying health conditions associated with COVID-19 severity^2^ may have been prone to collider bias, where the direction of effect measures changed based on model choice for some conditions. This was further underlined by higher testing and a seemingly protective effect when using the pseudo-test-negative design and true test-negative design for healthcare use variables that reversed when using the case-control design. Thus, comparisons of positivity rates require careful interpretation by examining the reasons for testing.^18^ In the context of low overall levels of testing, the case-control design may have mitigated some potential sources of collider bias, with the assumption that those not tested are similar to those who tested negative.^17,18^

Some underlying health conditions remained associated with diagnoses using the case-control design, reflecting either unmeasured confounding or possible biological susceptibility to infection if exposed.^10,11,21,53,54^ For example, dementia and frailty remained independently associated with diagnosis, potentially due to unmeasured confounding such as higher rates of contacts with care-givers or residence in other types of congregate settings such as retirement homes. Thus, underlying health conditions like dementia/frailty represent targets for prevention with strategies tailored to reduce exposures among individuals characterized by these individual determinants.

Over the study period, the testing criteria in Ontario shifted from returning symptomatic travellers, to severely symptomatic individuals and those with occupational exposure, to additional testing of asymptomatic individuals.^32-35,37^ These distinctions may have created differences in symptomatic tested individuals, compared to all tested individuals. In our study, the restriction of the test-negative design to symptomatic individuals did not yield substantially different results than the test-negative design including symptomatic and asymptomatic individuals for most determinants, but this may have been partly due to the high (74.6%) missingness of symptom information.

The independent association between high PM_2.5_ and diagnosis may reflect unmeasured social determinants of health.^55,56^ However, studies have also implicated environmental pollution as having a biological relationship to the risk and severity of COVID-19.^10,11,14^

We identified increased odds of diagnosis associated with household density, apartment building percentage, uncoupled status, essential work, educational attainment, and recent immigration, consistent with findings from other settings.^51,57,58^ Household size has been shown to be a consistent risk factor across a broad range of settings.^59,60^ These higher attack rates are likely due to prolonged and physically closer in-person contacts occurring more frequently within the household.^60^ Essential services and occupations have also been associated with higher exposure risk,^61^ either because such jobs cannot be done feasibly with proper protections, or because protective policies and materials are not issued, leaving workers at high risk.^62,63^

Higher percentages of recent immigrants in an area were associated with diagnosis, even after adjustment, although the percentage of visible minorities was not. Both variables might represent residual measures of structural racism, potentiating increased risk of SARS-CoV-2 exposure and COVID-19 severity,^64-66^ including COVID-19-related hospitalization/death.^9,17,29,58^ We found the association between visible minority status and diagnosis was attenuated after adjustment for individual, environmental, and other social determinants of health. These findings likely reflect what is already known about race and ethnicity as social constructs and social determinants of health.^67^ Finally, the fact that there was little variation in the odds of testing across levels of most of the social determinants suggests that testing resources may not be adequately prioritizing individuals at greatest risk.^68^

Taken together, our findings suggest a need to increase and/or redirect resources that specifically address social determinants such as household density^48,69^ (e.g., voluntary isolation centres^70^, wrap-around services^71^), occupational risk^62,66^ (e.g., paid sick leave^72^, workplace testing,^73^ improved ventilation^62^), and other mediators of structural racism^68,74,75^ (e.g. community-led outreach testing^76^). Our findings also suggest prioritizing COVID-19 vaccination strategies that reach communities and workplaces experiencing the highest rates of cases. Although there have been national recommendations for an equity lens to the public health response to COVID-19,^46^ much of the existing response on COVID-19 equity and outreach to marginalized communities has been accomplished through smaller, independent groups, including volunteer organizations.^77-79^

## Limitations

Our diagnoses were restricted to laboratory-confirmed cases and to the 88% of total provincial diagnoses that were available via OLIS, and thus may have missed probable cases as well as the remaining laboratory-confirmed cases that may have different determinants of infection. Results are also conditioned on the assumption that determinants remained constant across the study period, whereas surveillance data suggest shifts in how infections propagate between social networks.^80^ Future analyses include examining changes in the direction and magnitude of determinants over the course of the outbreak. Our models also adjusted for public health region, within which many social determinants cluster^51^, and thus we cannot infer from the results presented how social determinants of diagnosis may vary between and within these geographic regions. Social determinants were measured at the area level and were not available at the individual level; however, by describing individuals’ neighbourhoods, analyses reflect the role of structural and environmental determinants for individuals living amongst them. Fully-adjusted models in these analyses adjusted for a large number of covariates, potentially creating scenarios of over-adjustment. However, the directions of effect estimates generally remained the same after full adjustment, and the sample size of our analyses provided adequate statistical power. Finally, some important determinants, such as obesity,^23,81^ were not available for our study.^82^

## Conclusion

Individual-level risks for diagnosis defined by demographic and health-related determinants representing general targets of current response strategies appear to be subject to collider bias. Moving forward and advancing the response necessitates characterizing and addressing the social determinants potentiating heterogeneity in acquisition and transmission risks with risk-tailored, community-based interventions to reduce COVID-19 burden.

## Data Availability

The data referred to in the manuscript are private data concerning the health of all individuals living in Ontario, Canada. The data are shared in a secure manner between the Ontario Health Insurance Plan and ICES, a not-for-profit research institute. The de-identified and aggregated data may be available upon consultation with ICES.

## Disclaimers

The opinions, results, and conclusions reported in this paper are those of the authors and are independent from the funding sources. Parts of this material are based on data and/or information compiled and provided by the Canadian Institute for Health Information (CIHI) and by Cancer Care Ontario (CCO). However, the analyses, conclusions, opinions, and statement expressed herein are those of the authors, and not necessarily those of CIHI or CCO. No endorsement by ICES, MOHLTC, CIHI, or CCO is intended or should be inferred.

## Acknowledgements

We thank IQVIA Solutions Canada Inc. for use of their Drug Information File, as well as Owen Langman for technical assistance. Finally, we are grateful to the 14.7 million Ontario residents without whom this research would be impossible.

## Author contributions

JK, SM, and SB designed the study. ACa conducted all data analyses (dataset and variable creation and statistical modeling). JK, SM, SB, RK, ACa designed the analyses plans and conducted variable selection, with input from HC and AKC on variable selection and definitions. MAH, MD, LCR, TW contributed to analytic plans related to collider bias. BC contributed to data analyses and data preparation for the symptomatic dataset. MS prepared the figures. MS, JK, SB, and SM wrote the manuscript. All authors interpreted the data and critically reviewed and edited the manuscript.

