## Supplemental Table 1 for "The Individual and Social Determinants of COVID-19 Diagnosis in Ontario, Canada: A Population-Wide Study"

**Supplemental Table 1.** List and definitions of all covariates in the analyses.^a^

| **Variable** | **Definition** |
| --- | --- |
| Age | For individuals tested in OLIS, age was determined from their test record in OLIS. For individuals not tested in OLIS, age was determined from the Registered Persons Database. This variable was included *a priori* as hypothesized to be directly related to COVID-19 infection risk.^2^ |
| Sex | For individuals tested in OLIS, sex was determined from their test record in OLIS. For individuals not tested in OLIS, sex was determined from the Registered Persons Database. This variable was included *a priori* as hypothesized to be directly related to COVID-19 infection risk.^2^ |
| Rural residence | Defined using 2016 Census data as a residence that is not within the commuting zone of a population centre of greater than 10,000 people. Information about rural areas was matched to individuals using postal code. This variable was included as believed to be a marker for access to COVID-19 testing.^81^ |
| Public Health Region | Taken from Public Health Unit (PHU) information from the Registered Persons Database. Regions were defined as follows:  Central East: PHU 35 (Haliburton, Kawartha, Pine Ridge District Health Unit), 55 (Peterborough County—City Health Unit), 60 (Simcoe Muskoka District Health Unit)  Central West: PHU 27 (Brant County Health Unit), 34 (Haldimand-Norfolk Health Unit), 36 (Halton Regional Health Unit), 37 (City of Hamilton Health Unit), 46 (Niagara Regional Area Health Unit), 65 (Waterloo Health Unit), 66 (Wellington-Dufferin-Guelph Health Unit)  Durham: PHU 30 (Durham Regional Health Unit)  Eastern: PHU 38 (Hastings and Prince Edward Counties Health Unit), 41 (Kingston, Frontenac and Lennox and Addington Health Unit), 43 (Leeds, Grenville and Lanark District Health Unit), 57 (Renfrew County and District Health Unit), 58 (The Eastern Ontario Health Unit)  North: PHU 26 (The District of Algoma Health Unit), 47 (North Bay Parry Sound District Health Unit), 49 (Northwestern Health Unit), 56 (Porcupine Health Unit), 61 (Sudbury and District Health Unit), 62 (Thunder Bay District Health Unit), 63 (Timiskaming Health Unit)  Ottawa: PHU 51 (City of Ottawa Health Unit)  Peel: PHU 53 (Peel Regional Health Unit)  South West: PHU 31 (Elgin-St. Thomas), 33 (Grey Bruce Health Unit), 39 (Huron County Health Unit), 40 (Chatham-Kent Health Unit), 42 (Lambton Health Unit), 44 (Middlesex-London Health Unit), 52 (Oxford), 54 (Perth District Health Unit), 68 (Windsor-Essex County Health Unit)  Toronto: PHU 95 (City of Toronto Health Unit)  York: PHU 70 (York Regional Health Unit) |
| Asthma | An ICES-specific asthma database was used to identify patients with asthma, based on 2 or more ambulatory care visits and/or 1 or more hospitalizations. This variable was included *a priori* as hypothesized to be directly related to COVID-19 infection risk, as a result of its relationship to severe COVID-19 outcomes.^29^  OHIP  OHIP diagnostic code: 493  DAD  ICD-9 diagnostic code: 493  ICD-10 diagnostic codes: J45, J46 |
| Chronic obstructive pulmonary disease (COPD) | An ICES-specific COPD database was used to identify patients with COPD, based on 1 or more ambulatory care visits and/or 1 or more hospitalizations. The algorithm to identify COPD patients was only validated in those aged 35 years or older. This variable was included *a priori* as hypothesized to be directly related to COVID-19 infection risk.^30^  OHIP  OHIP diagnostic codes: 491, 492, 496  DAD  ICD-9 diagnostic codes: 491, 492, 496  ICD-10 diagnostic codes: J41, J42, J43, J44 |
| Hypertension | An ICES-specific hypertension database was used to identify patients with hypertension, based on 1 or more DAD diagnoses or 2 or more OHIP diagnoses in a two-year period; or 1 OHIP diagnosis followed by an OHIP/DAD diagnosis within two years. This variable was included *a priori* as hypothesized to be directly related to COVID-19 infection risk.^30^  DAD, SDS:  ICD-9 diagnostic codes: 401, 402, 403, 404, 405  ICD-10 diagnostic codes: I10, I11, I12, I13, I15  OHIP:  OHIP diagnostic codes: 401, 402, 403, 404, or 405 |
| Diabetes | An ICES-specific diabetes database was used to identify patients with diabetes, based on 2 OHIP diagnostic codes or 1 OHIP service code or 1 DAD admission within 2 years. This variable was included *a priori* as hypothesized to be directly related to COVID-19 infection risk.^30^  OHIP  OHIP diagnostic code: 250  OHIP service codes: Q040, K029, K030, K045, K046  DAD, SDS  ICD-9 diagnostic code: 250  ICD-10 diagnostic codes: E10, E11, E13, E14 |
| History of congestive heart failure (CHF) | An ICES-derived CHF database was used to identify patients with CHF, based on 1 NACRS, DAD, SDS, or OHIP claim and a second claim (from either) in 1 year. The CHF database is limited to those aged 40 years or older. This variable was included *a priori* as hypothesized to be directly related to COVID-19 infection risk.^30^  OHIP:  OHIP diagnostic code: 428  DAD, SDS:  ICD-9 diagnostic code: 428  ICD-10 diagnostic codes: I500, I501, I509 |
| Dementia/Frailty | Dementia (ICES cohort) definition:  1 hospitalization for dementia and/or 3 ambulatory visits for dementia, each separated by at least 30 days, within 2 years and/or 1 prescription from ODB. This variable was included *a priori* as hypothesized to be directly related to COVID-19 infection risk,^30^ as well as a marker for healthcare access, mobility, and household-level exposures.^39,82^  OHIP definition:  OHIP diagnostic codes: 290, 331  DAD, SDS definition:  ICD-9 diagnostic codes: 0461, 290.0, 290.1, 290.2, 290.3, 290.4, 294, 331.0, 331.1, 331.5  ICD-10 diagnostic codes: F00, F01, F02, F03, G30  ODB definition:  1 prescription for a cholinesterase inhibitor  Frailty:  Hospital Frailty Risk Score - Based on an algorithm already derived,^83^ using all DAD hospitalizations in the 5 years before index.  NOTE: Codes used in this risk score also include frailty-related conditions such as dementia and other chronic conditions that are reported separately |
| Cancer | We used the Ontario Cancer Registry to identify patients with any cancer diagnosed in the 5 years prior to index, except for non-melanoma skin cancer (ICD-O-3 Topography = C44 and Morphology = 87xx3). This variable was included *a priori* as hypothesized to be directly related to COVID-19 infection risk.^2^ |
| Chronic kidney disease (CKD) | This variable was included *a priori* as hypothesized to be directly related to COVID-19 infection risk.^30^ We defined this variable as having a CKD diagnosis code in DAD, NACRS, OHIP in the past 5 years, or:   - At least 1 dialysis code in each of the 3 months prior to index - Diagnosis and procedure codes found in concept dictionary   OHIP  OHIP diagnostic codes: 403, 585  NACRS, DAD  ICD-10 diagnostic codes: E102, E112, E132, E142, I12, I13, N08, N18, N19 |
|  | Patients who were on chronic dialysis in the year before index date, identified as those with at least 2 of any of the following codes in OHIP, DAD, or SDS separated by at least 90 days, but less than 150 days  OHIP  OHIP service codes: R849, G323, G325, G326, G860, G862, G865 G863, G866, G330, G331, G332, G333, G861, G082, G083, G085, G090, G091, G092, G093, G094, G095, G096, G294, G295, G864, H540, H740  DAD, SDS  CCI procedure codes: 5195, 6698  CCP procedure code: 1PZ21 |
| Immunocompromised (HIV, transplant, immunosuppressive therapy) | We included immunosuppressive conditions *a priori* as hypothesized to be directly related to COVID-19 infection risk.^30^  HIV  An ICES-specific HIV database was used to identify patients with HIV, based on 3 physician claims in 3 years with OHIP diagnostic codes: 042, 043 or 044  CORRLINK  CORRLINK is an ICES-specific database which links CORR and DAD data. This database only includes patients that have received an organ transplant and does not include dialysis patients.  History of allogenic/autologous bone marrow transplant (DAD, OHIP): We identified those who had a history of allogenic bone marrow transplant before index date using the following combination of diagnostic codes:  DAD:  Before 2002: CCP code (prcode) = 53.0  2002 and onwards: CCI code (incode) = 1WY19, 1LZ19HHU7, 1LZ19HHU8  OHIP:  Feecode = Z426  Any hospitalization (any diagnosis field) with the following codes:   - Sickle-cell disease (ICD10 D57.0 – D57.2; D57.8 OR ICD9 282.6); - Hereditary immunodeficiency (ICD10 D80-D84; D89.8, D89.9 and ICD9 279); - Neutropenia (ICD10 D70; ICD9 288.0); - Functional disorders of polymorphonuclear neutrophils & genetic anomalies of leukocytes (ICD10 D71-D72; ICD9 288.1, 288.2); - Hyposplenism, hypersplenism and chronic congestive splenomegaly (ICD10 D73.0, D73.1, D73.2; ICD9 289.4 or 289.5); - Asplenia (ICD9 759.0; ICD10 Q89.0); - HIV positive (ICD9 042; ICD10 B20) or identified in HIV database |
| Advanced liver disease (Cirrhosis or Decompensated Cirrhosis) | We included advanced liver disease *a priori* as hypothesized to be directly related to COVID-19 infection risk.^30^  Defined using the Cirrhosis Algorithm 9 [from ^84^]:  Two or more physician visits (diagnosis code 571), or  one or more hospital diagnosis of cirrhosis, using the following diagnostic codes:  ICD-9 : 456.1, 571.2, 571.5  ICD-10: I85.9, I98.2, K70.3,K71.7, K74.6  Defined using the Decompensated Cirrhosis Algorithm 5 (from above reference):  One or more physician visits with diagnosis code 571 and (one or more hospital diagnosis or one or more procedure), using the following diagnostic codes:  ICD-9: 456.0, 456.2, 572.2, 572.3, 572.4, 782.4, 789.5  ICD-10: I85.0, I86.4, I98.20, I98.3, K721, K729, K76.6, K76.7, R17, R18  CCI: 1.NA.13.BA-FA, 1.NA.13.BA-X7, 1.NA.13.BA-BD, 1.KQ.76GP-NR, 1.OT.52.HA  CCP: 1006, 6691  OHIP: J057, Z591 |
| History of cardiac ischemia | This variable was included *a priori* as hypothesized to be directly related to COVID-19 infection risk.^30^  Cardiac ischemic disease (DAD, SDS): Any comorbidity in the past 5 years (DAD, any diagnosis field) or history of procedure in past 20 years (DAD, SDS):  Comorbidity (DAD, any diagnosis in the past 5 years):  Angina: ICD-10 diagnostic codes: I20  ICD-9: 413  Chronic Ischemic Heart Disease: ICD-10 diagnostic codes: I25  ICD-9: 4140, 4148, 4149    Myocardial infarction: ICD-10 diagnostic codes: I21, I22  ICD-9: 410, 411, 412    Procedure (DAD & SDS):  Coronary Artery Bypass Grafting:  CCI procedure codes: 1IJ76  CCP procedure codes: 481    Percutaneous Coronary Intervention:  CCI procedure codes: 1IJ50, 1IJ5, 1IJ57  CCP procedure codes: 4802, 4803 |
| History of transient ischemic attack or acute ischemic stroke | This variable was included *a priori* as hypothesized to be directly related to COVID-19 infection risk.^30^  Transient Ischemic Attack:  DAD and NACRS were used to identify patients with a history of a transient ischemic attack, based on at least 1 hospitalization or ED visit with a diagnosis coded with one of the following codes:  ICD-9 diagnostic codes: 435, 3623  ICD-10 diagnostic codes: G450, G451, G452, G453, G458, G459, H340  Acute Ischemic Stroke:  DAD was used to identify patients with a history of acute ischemic stroke, based on at least 1 hospitalization with a main diagnosis coded with one of the following codes:  ICD-9 diagnostic codes: 43301, 43311 43321 43331 43381 43391 434, 436  ICD-10 diagnostic codes: I63, I64, H34.1 |
| Schizophrenia | This variable was included as believed to be a marker for access to COVID-19 testing, as well as a marker for other healthcare access.^85^  DAD/OMHRS: Hospitalizations for schizophrenia and related disorders identified using Mental Health and Addictions Scorecard and Evaluation Framework (MHASEF) diagnostic groupings for 'Schizophrenia Spectrum and Other Psychotic Disorders',^86^ using codes F20 (excluding F20.4), F22-F25, F28, F29, and F53.1.  DSM-IV or provisional diagnosis codes: 295.x (all 295 codes), 297.x (all 297 codes), 298.x (all 298 codes) or provisional diagnosis 5.  OHIP, NACRS: Ambulatory emergency department visits and physician office visits for schizophrenia identified with OHIP diagnosis code 295, or ICD-10 codes F20 or F25. |
| Substance abuse | This variable was included as believed to be a marker for access to COVID-19 testing, as well as a marker for other healthcare access.^87^  DAD/OMHRS: Hospitalizations for substance abuse and addictive disorders identified using Mental Health and Addictions Scorecard and Evaluation Framework (MHASEF) diagnostic groupings for 'Substance-related and Addictive Disorders’,^86^ using codes F55 and F10-F19.  DSM-IV or provisional diagnosis codes: 291.x (all 291 codes, excluding 291.82), 292.x (all 292 codes, excluding 292.85), 303.x (all 303 codes), 304.x (all 304 codes), 305.x (all 305 codes), or provisional diagnosis 4.  OHIP, NACRS: Ambulatory emergency department visits and physician office visits for substance abuse identified with OOHIPI diagnosis codes 303 and 304, or ICD-10 codes F55 or F1 |
| ADG quintiles | ADGs are clusters of similar diagnoses, defined based on their estimated impact on health services resource consumption. Membership in an ADG was calculated by the Johns Hopkins Adjusted Clinical Groups (ACG) software, ACG® System Version 10, based on DAD and OHIP records in the past 2 years. Individuals were ranked by quintile based on the number of ADGs they belonged to.  Number of ADGs assigned by Johns Hopkins Adjusted Clinical Groups (ACG) System Version 10 software, using DAD and OHIP records in the 2 years pre-index. We ranked individuals in the province by number of ADGs into 5 categories (quintiles), each containing approximately 20% of the overall Ontario population. There are a total of 32 ADGs that can be defined by the software: “Time Limited: Minor”, “Time Limited: Minor-Primary Infections”, “Time Limited: Major”, “Time Limited: Major-Primary Infections”, “Allergies”, “Asthma”, “Likely to Recur: Discrete”, “Likely to Recur: Discrete-Infections”, “Chronic Medical: Stable”, “Chronic Medical: Unstable”, “Chronic Speciality: Stable-Orthopedic”, “Chronic Specialty: Stable-Ear, Nose, Throat”, “Chronic Specialty: Stable-Eye”, “Chronic Specialty: Unstable-Orthopedic”, “Chronic Specialty: Unstable-Ear, Nose, Throat”, “Chronic Specialty: Unstable-Eye”, “Dermatologic”, “Injuries/Adverse Effects: Minor”, “Injuries/Adverse Effects: Major”, “Psychosocial: Time Limited, Minor”, “Psychosocial: Recurrent or Persistent, Stable”, “Psychosocial: Recurrent or Persistent, Unstable”, “Signs/Symptoms: Minor”, “Signs/Symptoms: Uncertain”, “Signs/Symptoms: Major”, “Discretionary”, “See and Reassure”, “Prevention/Administrative”, “Malignancy”, “Pregnancy”, and “Dental”. More information can be found in the following reference: ^42^. |
| Hospital admissions, last 3 years | DAD/OMHRS: Number of acute-care hospital admissions in the 3 years before the index date. |
| Outpatient visits, last year | OHIP: Number of physician office, long-term care, or home visits in the 1 year before the index date. |
| Influenza vaccine received, 2019-2020 season | An OHIP billing with any of the following fee codes from October 1, 2019 up to 14 days before the index date: G590, G591, G592, Q130, Q590, Q690, Q691;  or, an ODB billing with any of the following Drug Identification Numbers (DINs) from October 1, 2019 up to 14 days before the index date: 02015986, 02223929, 02269562, 02346850, 02362384, 02365936, 02420643, 02420686, 02420783, 02426544, 02428881, 02432730, 02473283, 09857501 |
| PM_2.5_ | 3-year average for 2014-2016 in a 1km x 1km area, from satellite observations of aerosol optical depth in combination with outputs from a global atmospheric chemistry transport model.^88^ The PM_2.5_ estimates were further calibrated using the information on land cover, elevation, and aerosol composition, producing mean concentration of PM_2.5_ (1×1km) yearly between 2000 and 2016.^43^ Covering all of North America including Ontario, these annual estimates of PM_2.5_ closely agree with ground-level measurements at fixed-site monitors across North America (R^2^ =0.82, n = 1440). To temporally align air pollution exposures with our study period, we derived three-year moving average estimates for PM_2.5_ and NO_2_ between 2014 and 2016, and thus yielding historical exposures to these two common air pollutants in our study cohort. PM_2.5_ levels were attributed to subjects at the geographic level of postal code. |
| NO_2_ | 3-year average for 2014-2016 (30m x 30m) from a national land-use regression model developed using NO_2_ observations at fixed-site monitors across Canada.^89^ The model explains 73% of the variability in annual 2006 measurements of NO_2_.^44^ This NO_2_ land-use regression model has recently been updated using more recent NO_2_ monitoring data and land-use and vehicular predictors, thereby covering a more recent period of 2014 to 2016. To temporally align air pollution exposures with our study period, we derived three-year moving average estimates for PM_2.5_ and NO_2_ between 2014 and 2016, and thus yielding historical exposures to these two common air pollutants in our study cohort. NO_2_ levels were attributed to subjects at the geographic level of postal code. |
| Dissemination Area (DA) | A dissemination area (DA) is the smallest standard geographic area for which all census data are disseminated. A DA generally comprises approximately 400-700 people, but in densely populated cities may contain several thousand people. DAs cover all the territory of Canada.^49^  We assigned subjects to a DA using postal code, as recorded in OLIS. For those who did not have a valid postal code recorded or were not tested for COVID-19, we used the individual’s postal code as listed in the Registered Persons Database. |
| Household density quintile | Average number of persons in private households, calculated at the DA level using the 2016 Census data.^94^ DAs across the province were ranked by average number of persons per household into 5 categories (quintiles), such that each group contained approximately one-fifth of the DAs. |
| Apartment building density grouping | Calculated at the DA level using 2016 Census data.^95^ This variable was computed by identifying the number of individuals reporting living in an “apartment in a building that has five or more storeys” or “apartment in a building that has fewer than five storeys” and dividing this value by the total number of individuals having answered questions about occupied private dwellings by structural type of dwelling. This yielded a percentage of dwellings in each DA considered to be apartment buildings. DAs across the province were then ranked by these percentages into three groups with cut-offs at the 60th and 80th percentiles, due to a zero-inflated distribution of DAs. |
| Uncoupled quintile | Calculated at the DA level using 2016 Census data.^93^ “Uncoupled” individuals are individuals who, in the 2016 Census, reported having the following relationship statuses (wording from 2016 Census): never married (persons who have never legally married and are not living with a person as a couple); separated (persons who are married but who are no longer living with their spouse [for reasons other than, for example, illness, work or school], have not obtained a divorce and are not living with a person as a couple; divorced (persons who have obtained a legal divorce, have not remarried and are not living with a person as a couple); and widowed (persons who have lost their married spouse through death, have not remarried and are not living with a person as a couple).  The number of individuals fulfilling this description in a given DA was divided by the total number of individuals with marital status in the DA, yielding a percentage figure for each DA. DAs across the province were then ranked by these percentages into quintiles, with the lowest 1/5 of DAs comprising the first quintile, and so on. |
| Essential work quintile | Calculated at the DA level, using 2016 Census data.^97^ For each DA, we calculated the number of individuals ≥15 years old that were working in one of the following Census-defined work categories: Sales and service occupations; trades, transport and equipment operators and related occupations; natural resources, agriculture and related production occupations; and occupations in manufacturing and utilities.  DAs across the province were then ranked by these percentages into quintiles, with the lowest 1/5 of DAs comprising the first quintile, and so on. |
| Household income quintile | Calculated at the DA level using 2016 Census data^90^ by multiplying the median income (before-tax) by the number of households and dividing by the sum of single-person equivalent to obtain income per single person equivalent. For DAs where median income was unavailable, neighbouring DAs were used to estimate income per single person equivalent. DA-based income quintiles were constructed separately for each census metropolitan area or census agglomeration (one or more adjacent municipalities integrated via commuting flows). DAs within each such area were ranked from the lowest average income per single-person equivalent to the highest, and DAs were assigned to five groups, such that each group contained approximately one-fifth the total in-scope population of each area. |
| Limited educational attainment quintile | Calculated at the DA level using 2016 Census data.^92^ This variable was computed by identifying the number of adults aged 25-64 reporting having “no certificate, diploma, or degree” and dividing this value by the total number of individuals aged 25-64 having answered questions about their highest certificate, diploma or degree. This yielded a percentage of individuals in each DA considered to have no certificate, diploma, or degree. DAs across the province were then ranked by these percentages into quintiles, with the lowest 1/5 of DAs comprising the first quintile, and so on. |
| Visible minority quintile | Calculated at the DA level, using 2016 Census data.^97^ An individual was marked as “self-identify as a visible minority” if they reported being one or more of the following (wording from the 2016 Census): “South Asian (e.g., East Indian, Pakistani, Sri Lankan, etc.), Chinese, Black, Filipino, Latin American, Arab, Southeast Asian (e.g., Vietnamese, Cambodian, Laotian, Thai, etc.), West Asian (e.g., Iranian, Afghan, etc.), Korean, Japanese, or Other—specify”.  DAs across the province were then ranked by these percentages into quintiles, with the lowest 1/5 of DAs comprising the first quintile, and so on. |
| Recent immigration grouping | Calculated at the DA level, using 2016 Census data.^96^ This value was obtained by identifying the number of individuals in a DA that reported having immigrated in the past 5 years, and dividing this number by the number of individuals in the DA who reported their immigration status over the past five years (i.e., were immigrants or non-immigrants, regardless of the time period of immigration).  DAs across the province were then ranked by percentage of recent percentages into three groups with cut-offs at the 60th and 80th percentiles, due to a zero-inflated distribution of DAs. |

^a^ OHIP=Ontario Health Insurance Plan, DAD=Discharge Abstract Database, NACRS=National Ambulatory Care Reporting System, SDS=Same Day Surgery, CCI=Canadian Classification of Health Interventions, CCP=Canadian Classification of Procedures, CORR=Canadian Organ Replacement Register, ORRS=Ontario Renal Reporting System, ODD=Ontario Diabetes Database, OCR=Ontario Cancer Registry, OMID=Ontario Myocardial Infarction Database, CHF=Ontario Congestive Heart Failure Database, ODB=Ontario Drug Benefit, HIV=Ontario HIV database
