## Supplemental Table 2 for "The Individual and Social Determinants of COVID-19 Diagnosis in Ontario, Canada: A Population-Wide Study"

**Supplemental Table 2**. Characteristics of study population tested and not tested for SARS-CoV-2 in Ontario between March 1, 2020 and June 20, 2020.

|  | **Tested individuals**  **N = 758,691** | | **Symptomatic tested individuals only**  **N = 89,408** | | **All individuals**  **N = 14,695,579** | |
| --- | --- | --- | --- | --- | --- | --- |
|  | **Tested negative**  **(N = 733,661)** | **Tested positive**  **(N = 25,030)** | **Tested negative and symptomatic**  **(N = 85,355)** | **Tested positive and symptomatic (N = 4,053)** | **Untested control group**  **(N = 13,936,888)** | **All tested**  **(N =758,691)** |
| **Demographic characteristics** |  |  |  |  |  |  |
| Age: mean ± SD | 49.52 ± 20.99 | 47.45 ± 19.77 | 47.91 ± 20.80 | 50.83 ± 20.39 | 40.31 ± 22.91 | 49.45 ± 20.95 |
| **Age groups (years):**  **N (%)** |  |  |  |  |  |  |
| 0-4 | 10,870 (1.5%) | 223 (0.9%) | 1,627 (1.9%) | 17 (0.4%) | 727,305 (5.2%) | 11,093 (1.5%) |
| 5-19 | 33,642 (4.6%) | 1,149 (4.6%) | 4,017 (4.7%) | 125 (3.1%) | 2,395,518 (17.2%) | 34,791 (4.6%) |
| 20-34 | 157,086 (21.4%) | 6,015 (24.0%) | 19,431 (22.8%) | 864 (21.3%) | 2,810,230 (20.2%) | 163,101 (21.5%) |
| 35-49 | 160,506 (21.9%) | 6,084 (24.3%) | 20,495 (24.0%) | 934 (23.0%) | 2,772,204 (19.9%) | 166,590 (22.0%) |
| 50-64 | 192,178 (26.2%) | 7,009 (28.0%) | 21,329 (25.0%) | 1,159 (28.6%) | 2,865,926 (20.6%) | 199,187 (26.3%) |
| 65-74 | 84,077 (11.5%) | 2,072 (8.3%) | 8,765 (10.3%) | 386 (9.5%) | 1,394,043 (10.0%) | 86,149 (11.4%) |
| 75-84 | 52,655 (7.2%) | 1,284 (5.1%) | 5,181 (6.1%) | 251 (6.2%) | 713,639 (5.1%) | 53,939 (7.1%) |
| ≥85 | 42,647 (5.8%) | 1,194 (4.8%) | 4,510 (5.3%) | 317 (7.8%) | 258,023 (1.9%) | 43,841 (5.8%) |
| Male sex: N (%) | 289,769 (39.5%) | 11,627 (46.5%) | 33,078 (38.8%) | 1,898 (46.8%) | 6,930,233 (49.7%) | 301,396 (39.7%) |
| Living in rural/small town: N (%)^a^ | 82,034 (11.2%) | 887 (3.5%) | 14,140 (16.6%) | 304 (7.5%) | 1,390,426 (10.0%) | 82,921 (10.9%) |
| **Chronic health conditions: N (%)** |  |  |  |  |  |  |
| Asthma | 136,556 (18.6%) | 3,887 (15.5%) | 17,831 (20.9%) | 671 (16.6%) | 2,085,118 (15.0%) | 140,443 (18.5%) |
| COPD | 34,261 (4.7%) | 715 (2.9%) | 4,616 (5.4%) | 168 (4.1%) | 249,636 (1.8%) | 34,976 (4.6%) |
| Hypertension | 226,111 (30.8%) | 7,488 (29.9%) | 24,364 (28.5%) | 1,313 (32.4%) | 2,836,790 (20.4%) | 233,599 (30.8%) |
| Diabetes | 108,758 (14.8%) | 4,268 (17.1%) | 11,763 (13.8%) | 684 (16.9%) | 1,363,667 (9.8%) | 113,026 (14.9%) |
| Congestive heart failure | 35,947 (4.9%) | 983 (3.9%) | 3,864 (4.5%) | 194 (4.8%) | 225,153 (1.6%) | 36,930 (4.9%) |
| Dementia or frailty | 31,804 (4.3%) | 1,106 (4.4%) | 3,399 (4.0%) | 296 (7.3%) | 124,295 (0.9%) | 32,910 (4.3%) |
| Cancer^b^ | 24,333 (3.3%) | 484 (1.9%) | 2,333 (2.7%) | 89 (2.2%) | 213,036 (1.5%) | 24,817 (3.3%) |
| Chronic kidney disease^b^ | 37,630 (5.1%) | 1,122 (4.5%) | 3,581 (4.2%) | 199 (4.9%) | 254,541 (1.8%) | 38,752 (5.1%) |
| Immunocompromised^c^ | 13,325 (1.8%) | 319 (1.3%) | 1,496 (1.8%) | 57 (1.4%) | 107,824 (0.8%) | 13,644 (1.8%) |
| Advanced liver disease | 9,020 (1.2%) | 212 (0.8%) | 1,045 (1.2%) | 35 (0.9%) | 78,747 (0.6%) | 9,232 (1.2%) |
| Ischemic heart disease | 33,855 (4.6%) | 880 (3.5%) | 3,773 (4.4%) | 168 (4.1%) | 324,682 (2.3%) | 34,735 (4.6%) |
| Ischemic stroke or transient ischemic attack^d^ | 13,777 (1.9%) | 411 (1.6%) | 1,531 (1.8%) | 77 (1.9%) | 99,503 (0.7%) | 14,188 (1.9%) |
| Schizophrenia^e^ | 7,526 (1.0%) | 220 (0.9%) | 732 (0.9%) | 42 (1.0%) | 64,494 (0.5%) | 7,746 (1.0%) |
| Substance abuse^e^ | 22,238 (3.0%) | 478 (1.9%) | 3,001 (3.5%) | 59 (1.5%) | 216,156 (1.6%) | 22,716 (3.0%) |
| **Healthcare use: N (%)** |  |  |  |  |  |  |
| **Adjusted Diagnostic Group quintile** |  |  |  |  |  |  |
| 1 (0 ADGs) | 31,959 (4.4%) | 1,571 (6.3%) | 3,006 (3.5%) | 247 (6.1%) | 1,616,852 (11.6%) | 33,530 (4.4%) |
| 2 (1-2 ADGs) | 101,895 (13.9%) | 3,601 (14.4%) | 11,419 (13.4%) | 595 (14.7%) | 3,150,969 (22.6%) | 105,496 (13.9%) |
| 3 (3-4 ADGs) | 149,173 (20.3%) | 5,101 (20.4%) | 17,220 (20.2%) | 782 (19.3%) | 3,385,602 (24.3%) | 154,274 (20.3%) |
| 4 (5-6 ADGs) | 150,394 (20.5%) | 5,115 (20.4%) | 17,736 (20.8%) | 822 (20.3%) | 2,626,403 (18.8%) | 155,509 (20.5%) |
| 5 (7-27 ADGs) | 300,240 (40.9%) | 9,642 (38.5%) | 35,974 (42.1%) | 1,607 (39.6%) | 3,157,062 (22.7%) | 309,882 (40.8%) |
| Hospital admissions, past 3 years: Mean (SD) | 0.44 ± 1.34 | 0.33 ± 1.21 | 0.45 ± 1.29 | 0.36 ± 1.09 | 0.19 ± 0.60 | 0.44 ± 1.34 |
| Outpatient physician visits, past year: Mean (SD) | 7.35 ± 8.68 | 7.06 ± 8.33 | 7.15 ± 8.40 | 7.07 ± 8.64 | 4.67 ± 6.05 | 7.34 ± 8.67 |
| Influenza vaccination (2019-2020 season) | 213,722 (29.1%) | 5,547 (22.2%) | 23,429 (27.4%) | 1,019 (25.1%) | 2,978,472 (21.4%) | 219,269 (28.9%) |
| **Environmental determinants^f^: N (%)** |  |  |  |  |  |  |
| **PM_2.5_ (µg/m^3^ yearly)** |  |  |  |  |  |  |
| 2 to <6 | 161,300 (22.0%) | 1,831 (7.3%) | 27,764 (32.5%) | 556 (13.7%) | 2,481,201 (17.8%) | 163,131 (21.5%) |
| 6 to <7 | 91,134 (12.4%) | 1,766 (7.1%) | 10,930 (12.8%) | 318 (7.8%) | 1,555,790 (11.2%) | 92,900 (12.2%) |
| 7 to <8 | 207,966 (28.3%) | 8,476 (33.9%) | 21,183 (24.8%) | 1,281 (31.6%) | 4,450,218 (31.9%) | 216,442 (28.5%) |
| 8 to <9 | 211,861 (28.9%) | 11,127 (44.5%) | 19,639 (23.0%) | 1,363 (33.6%) | 4,258,069 (30.6%) | 222,988 (29.4%) |
| ≥9 | 59,674 (8.1%) | 1,747 (7.0%) | 5,749 (6.7%) | 517 (12.8%) | 1,073,665 (7.7%) | 61,421 (8.1%) |
| **NO_2_ (ppb yearly)** |  |  |  |  |  |  |
| 0 to 6 | 328,613 (44.8%) | 5,237 (20.9%) | 48,579 (56.9%) | 1,500 (37.0%) | 5,505,976 (39.5%) | 333,850 (44.0%) |
| 6 to 8 | 170,693 (23.3%) | 5,599 (22.4%) | 17,477 (20.5%) | 1,079 (26.6%) | 3,409,506 (24.5%) | 176,292 (23.2%) |
| ≥8 | 232,629 (31.7%) | 14,111 (56.4%) | 19,209 (22.5%) | 1,456 (35.9%) | 4,903,460 (35.2%) | 246,740 (32.5%) |
| **Social determinants of health^g^ (area-level): N (%)** |  |  |  |  |  |  |
| **Household density quintile^h^** |  |  |  |  |  |  |
| 1 (0 – 2.1) | 162,623 (22.2%) | 3,639 (14.5%) | 20,066 (23.5%) | 672 (16.6%) | 2,474,391 (17.8%) | 166,262 (21.9%) |
| 2 (2.2 - 2.4) | 140,653 (19.2%) | 3,104 (12.4%) | 19,406 (22.7%) | 662 (16.3%) | 2,368,013 (17.0%) | 143,757 (18.9%) |
| 3 (2.5 - 2.6) | 104,896 (14.3%) | 2,721 (10.9%) | 13,059 (15.3%) | 531 (13.1%) | 1,866,317 (13.4%) | 107,617 (14.2%) |
| 4 (2.7 - 3) | 166,089 (22.6%) | 6,321 (25.3%) | 17,843 (20.9%) | 1,096 (27.0%) | 3,291,097 (23.6%) | 172,410 (22.7%) |
| 5 (3.1 - 5.7) | 152,578 (20.8%) | 8,929 (35.7%) | 14,345 (16.8%) | 1,024 (25.3%) | 3,775,489 (27.1%) | 161,507 (21.3%) |
| **Apartment building density category^i^** |  |  |  |  |  |  |
| 1 (0 – 7.3%) | 391,477 (53.4%) | 12,377 (49.4%) | 46,693 (54.7%) | 2,243 (55.3%) | 7,994,323 (57.4%) | 403,854 (53.2%) |
| 2 (7.4 - 37.7%) | 145,108 (19.8%) | 3,874 (15.5%) | 19,348 (22.7%) | 739 (18.2%) | 2,478,855 (17.8%) | 148,982 (19.6%) |
| 3 (37.7 - 104%) | 190,244 (25.9%) | 8,463 (33.8%) | 18,678 (21.9%) | 1,003 (24.7%) | 3,301,839 (23.7%) | 198,707 (26.2%) |
| **Uncoupled quintile^j^** |  |  |  |  |  |  |
| 1 (11.2 - 33.7%) | 150,044 (20.5%) | 3,539 (14.1%) | 20,243 (23.7%) | 824 (20.3%) | 3,130,907 (22.5%) | 153,583 (20.2%) |
| 2 (33.7 - 38.4%) | 128,561 (17.5%) | 3,915 (15.6%) | 15,262 (17.9%) | 779 (19.2%) | 2,698,847 (19.4%) | 132,476 (17.5%) |
| 3 (38.5 - 43.6%) | 127,689 (17.4%) | 4,632 (18.5%) | 14,539 (17.0%) | 693 (17.1%) | 2,579,005 (18.5%) | 132,321 (17.4%) |
| 4 (43.6 - 51.0%) | 145,560 (19.8%) | 5,591 (22.3%) | 15,894 (18.6%) | 713 (17.6%) | 2,633,710 (18.9%) | 151,151 (19.9%) |
| 5 (51.0 - 94.6%) | 174,985 (23.9%) | 7,037 (28.1%) | 18,781 (22.0%) | 976 (24.1%) | 2,732,838 (19.6%) | 182,022 (24.0%) |
| **Essential work quintile^k^** |  |  |  |  |  |  |
| 1 (0 – 32.5%) | 145,517 (19.8%) | 3,941 (15.7%) | 14,783 (17.3%) | 688 (17.0%) | 2,969,492 (21.3%) | 149,458 (19.7%) |
| 2 (32.5 – 42.3%) | 155,189 (21.2%) | 5,077 (20.3%) | 17,911 (21.0%) | 918 (22.6%) | 3,084,781 (22.1%) | 160,266 (21.1%) |
| 3 (42.3 – 49.8%) | 149,589 (20.4%) | 4,686 (18.7%) | 17,865 (20.9%) | 785 (19.4%) | 2,717,510 (19.5%) | 154,275 (20.3%) |
| 4 (50.0 – 57.5%) | 143,965 (19.6%) | 5,236 (20.9%) | 17,677 (20.7%) | 778 (19.2%) | 2,615,078 (18.8%) | 149,201 (19.7%) |
| 5 (57.5 – 114.3%) | 133,601 (18.2%) | 5,816 (23.2%) | 16,638 (19.5%) | 839 (20.7%) | 2,399,120 (17.2%) | 139,417 (18.4%) |
| **Household income quintile^l^** |  |  |  |  |  |  |
| 1 (lowest income) | 156,320 (21.3%) | 7,000 (28.0%) | 17,159 (20.1%) | 857 (21.1%) | 2,679,780 (19.2%) | 163,320 (21.5%) |
| 2 | 148,687 (20.3%) | 5,288 (21.1%) | 17,057 (20.0%) | 757 (18.7%) | 2,698,807 (19.4%) | 153,975 (20.3%) |
| 3 | 145,317 (19.8%) | 5,084 (20.3%) | 16,771 (19.6%) | 756 (18.7%) | 2,791,340 (20.0%) | 150,401 (19.8%) |
| 4 | 140,352 (19.1%) | 4,019 (16.1%) | 17,162 (20.1%) | 730 (18.0%) | 2,809,529 (20.2%) | 144,371 (19.0%) |
| 5 (highest income) | 138,103 (18.8%) | 3,419 (13.7%) | 16,822 (19.7%) | 910 (22.5%) | 2,815,238 (20.2%) | 141,522 (18.7%) |
| **Limited educational attainment quintile^m^** |  |  |  |  |  |  |
| 1 (0.0 - 4.1%) | 144,457 (19.7%) | 3,903 (15.6%) | 15,245 (17.9%) | 709 (17.5%) | 2,926,432 (21.0%) | 148,360 (19.6%) |
| 2 (4.1 - 7.5%) | 154,215 (21.0%) | 4,477 (17.9%) | 17,690 (20.7%) | 869 (21.4%) | 2,978,444 (21.4%) | 158,692 (20.9%) |
| 3 (7.5 - 11.4%) | 151,457 (20.6%) | 5,052 (20.2%) | 17,765 (20.8%) | 759 (18.7%) | 2,888,525 (20.7%) | 156,509 (20.6%) |
| 4 (11.4 - 17.1%) | 148,159 (20.2%) | 5,314 (21.2%) | 18,014 (21.1%) | 847 (20.9%) | 2,659,544 (19.1%) | 153,473 (20.2%) |
| 5 (17.1 - 94.3%) | 129,580 (17.7%) | 6,010 (24.0%) | 16,160 (18.9%) | 824 (20.3%) | 2,333,134 (16.7%) | 135,590 (17.9%) |
| **Visible minority quintile^n^** |  |  |  |  |  |  |
| 1 (0.0 - 2.2%) | 130,912 (17.8%) | 1,716 (6.9%) | 21,800 (25.5%) | 555 (13.7%) | 2,115,641 (15.2%) | 132,628 (17.5%) |
| 2 (2.2 - 7.5%) | 137,826 (18.8%) | 2,233 (8.9%) | 19,869 (23.3%) | 677 (16.7%) | 2,255,245 (16.2%) | 140,059 (18.5%) |
| 3 (7.5 - 18.7%) | 137,744 (18.8%) | 3,201 (12.8%) | 16,345 (19.1%) | 792 (19.5%) | 2,451,335 (17.6%) | 140,945 (18.6%) |
| 4 (18.7 - 43.5%) | 153,503 (20.9%) | 5,466 (21.8%) | 13,581 (15.9%) | 887 (21.9%) | 3,023,752 (21.7%) | 158,969 (21.0%) |
| 5 (43.5 - 102%) | 167,893 (22.9%) | 12,140 (48.5%) | 13,280 (15.6%) | 1,097 (27.1%) | 3,940,245 (28.3%) | 180,033 (23.7%) |
| **Recent immigration category^o^** |  |  |  |  |  |  |
| 1 (0.0 - 2.1%) | 401,300 (54.7%) | 8,271 (33.0%) | 54,845 (64.3%) | 2,002 (49.4%) | 6,967,468 (50.0%) | 409,571 (54.0%) |
| 2 (2.1 - 4.7% | 146,772 (20.0%) | 5,409 (21.6%) | 14,834 (17.4%) | 852 (21.0%) | 2,858,064 (20.5%) | 152,181 (20.1%) |
| 3 (4.7 - 41.2%) | 174,829 (23.8%) | 11,018 (44.0%) | 14,393 (16.9%) | 1,133 (28.0%) | 3,898,463 (28.0%) | 185,847 (24.5%) |

^a^ “Rural” defined as located outside the commuting zone of a city with greater than 10,000 population.

^b^ Participants were counted if they had a diagnosis in the last 5 years.

^c^ Participants considered “immunocompromised” if they were HIV positive, or had an organ or bone marrow transplant, or another immunodeficient condition.

^d^ This category includes individuals with ischemic stroke or transient ischemic attack in the last 20 years.

^e^ This category includes individuals with a diagnosis in the last 2 years.

^f^ Values of PM_2.5_ > 12µg/m^3^ or NO_2_ > 53ppb yearly has been found to be associated with increased risk of other respiratory illnesses.^80^ PM_2.5_ and NO_2_ values are provided at the postal code, not DA, level.

^g^ All variables in this category are area-level variables at the level of the Census Dissemination Area (DA).

^h^ Range of persons per dwelling.

^i^ The census counts are randomly rounded up or down to the nearest number divisible by 5, which causes some minor imprecision. 7.3% represents the 60^th^ percentile.

^j^ “Uncoupled” individuals are those never married (persons who have never legally married and are not living with a person as a couple); separated (persons who are married but who are no longer living with their spouse [for reasons other than, for example, illness, work or school], have not obtained a divorce and are not living with a person as a couple; divorced (persons who have obtained a legal divorce, have not remarried and are not living with a person as a couple); and widowed (persons who have lost their married spouse through death, have not remarried and are not living with a person as a couple).

^k^ Percentage of individuals in the area working in the following occupations: Sales and service occupations; trades, transport and equipment operators and related occupations; natural resources, agriculture and related production occupations; and occupations in manufacturing and utilities. Census counts for individuals are randomly rounded up or down to the nearest number divisible by 5, which causes some minor imprecision.

^l^ Income quintile has variable cut-off values in each city or census area, to take cost of living into account. A DA being in quintile 1 means it is among the lowest 20% DAs in its city by income.

^m^ Percentage of adults aged 25-64 in the area not having received any diploma.

^n^ Percentage of individuals in the area self-identifying as a visible minority. Census counts for individuals are randomly rounded up or down to the nearest number divisible by 5, which causes some minor imprecision.

^o^ Percentage of individuals in the area who are recent immigrants; 2.1% represents the 60^th^ percentile.
