## Supplemental Table 3 for "The Individual and Social Determinants of COVID-19 Diagnosis in Ontario, Canada: A Population-Wide Study"

**Supplemental Table 3.** Unadjusted, age/sex-adjusted, and fully-adjusted model results describing the odds of being tested for SARS-CoV-2 in Ontario, Canada between March 1 and June 20, 2020.

| **Demographic characteristics** | **Odds of testing (unadjusted)** | **Odds of testing (age/sex-adjusted)** | **Odds of testing (fully-adjusted)^a^** |
| --- | --- | --- | --- |
| **Sample size** | 14,695,579 | 14,695,579 | 14,695,579 |
| **Age groups (years) (ref: 0 – 4)** |  |  |  |
| 5-19 | 0.95 (0.93, 0.97) | 0.95 (0.93, 0.97) | 1.15 (1.13, 1.18) |
| 20-34 | 3.81 (3.73, 3.88) | 3.79 (3.72, 3.87) | 4.13 (4.05, 4.22) |
| 35-49 | 3.94 (3.86, 4.02) | 3.91 (3.83, 3.99) | 4.13 (4.05, 4.21) |
| 50-64 | 4.56 (4.47, 4.65) | 4.53 (4.44, 4.62) | 4.21 (4.12, 4.29) |
| 65-74 | 4.05 (3.97, 4.13) | 4.00 (3.92, 4.08) | 3.04 (2.98, 3.11) |
| 75-84 | 4.96 (4.85, 5.06) | 4.86 (4.76, 4.97) | 3.01 (2.95, 3.08) |
| ≥85 | 11.14 (10.91, 11.38) | 10.69 (10.46, 10.92) | 5.60 (5.47, 5.73) |
| Male sex | 0.67 (0.66, 0.67) | 0.69 (0.68, 0.69) | 0.76 (0.76, 0.76) |
| Living in rural/small town^b^ | 1.11 (1.10, 1.12) | 1.10 (1.09, 1.10) | 0.97 (0.96, 0.98) |
| **Underlying chronic health conditions** |  |  |  |
| Asthma | 1.29 (1.28, 1.30) | 1.31 (1.30, 1.31) | 1.09 (1.09, 1.10) |
| COPD | 2.65 (2.62, 2.68) | 1.91 (1.88, 1.93) | 1.23 (1.21, 1.24) |
| Hypertension | 1.74 (1.73, 1.75) | 1.25 (1.24, 1.26) | 0.98 (0.98, 0.99) |
| Diabetes | 1.61 (1.60, 1.62) | 1.22 (1.22, 1.23) | 0.98 (0.97, 0.99) |
| Congestive heart failure | 3.12 (3.08, 3.15) | 2.08 (2.05, 2.10) | 1.26 (1.24, 1.28) |
| Dementia or frailty score >15 | 5.04 (4.98, 5.10) | 3.15 (3.11, 3.19) | 2.12 (2.09, 2.15) |
| Cancer^c^ | 2.18 (2.15, 2.21) | 1.64 (1.62, 1.67) | 1.13 (1.12, 1.15) |
| Chronic kidney disease^c^ | 2.89 (2.86, 2.93) | 2.10 (2.08, 2.13) | 1.31 (1.29, 1.32) |
| Immunocompromised^d^ | 2.35 (2.31, 2.39) | 2.11 (2.07, 2.14) | 1.30 (1.28, 1.33) |
| Advanced liver disease | 2.17 (2.12, 2.22) | 1.78 (1.74, 1.82) | 1.14 (1.11, 1.16) |
| Ischemic heart disease | 2.01 (1.99, 2.03) | 1.56 (1.55, 1.58) | 1.02 (1.01, 1.03) |
| Ischemic stroke or transient ischemic attack^e^ | 2.65 (2.60, 2.70) | 1.77 (1.73, 1.80) | 1.15 (1.13, 1.17) |
| Schizophrenia^f^ | 2.22 (2.17, 2.27) | 2.03 (1.98, 2.08) | 1.24 (1.21, 1.27) |
| Substance abuse^f^ | 1.96 (1.93, 1.99) | 1.90 (1.88, 1.93) | 1.17 (1.16, 1.19) |
| **Healthcare use** |  |  |  |
| **Aggregated Diagnostic Group quintile (ref: 0 ADGs)** |  |  |  |
| 2 (1-2 ADGs) | 1.61 (1.59, 1.63) | 1.72 (1.70, 1.74) | 1.64 (1.62, 1.66) |
| 3 (3-4 ADGs) | 2.20 (2.17, 2.22) | 2.24 (2.21, 2.27) | 2.07 (2.04, 2.10) |
| 4 (5-6 ADGs) | 2.86 (2.82, 2.89) | 2.73 (2.70, 2.76) | 2.44 (2.41, 2.48) |
| 5 (7-27 ADGs) | 4.73 (4.68, 4.79) | 4.04 (4.00, 4.09) | 3.11 (3.06, 3.15) |
| **Hospital admissions, past 3 years (ref: 0 admissions)** |  |  |  |
| 1 | 1.38 (1.37, 1.39) | 1.40 (1.39, 1.41) | 0.99 (0.98, 1.00) |
| 2 | 2.26 (2.23, 2.28) | 1.98 (1.96, 2.01) | 1.16 (1.14, 1.17) |
| ≥3 | 4.90 (4.84, 4.96) | 4.08 (4.03, 4.13) | 1.75 (1.73, 1.78) |
| **Outpatient physician visits, past year (ref: 0 – 1 visits)** |  |  |  |
| 2 – 4 | 1.55 (1.54, 1.56) | 1.48 (1.47, 1.49) | 1.08 (1.07, 1.09) |
| 5 – 8 | 2.00 (1.99, 2.02) | 1.78 (1.77, 1.79) | 1.11 (1.10, 1.12) |
| 9 – 14 | 2.57 (2.55, 2.59) | 2.13 (2.11, 2.15) | 1.17 (1.16, 1.18) |
| ≥15 | 3.70 (3.67, 3.73) | 2.85 (2.82, 2.87) | 1.36 (1.34, 1.37) |
| Influenza vaccination (2019-2020 season) | 1.50 (1.49, 1.50) | 1.26 (1.25, 1.27) | 1.08 (1.08, 1.09) |
| **Environmental determinants^g^** |  |  |  |
| **PM_2.5_ category (µg/m^3^) (ref: 2 – 6)** |  |  |  |
| 6-7 | 0.91 (0.90, 0.92) | 0.91 (0.90, 0.91) | 0.97 (0.97, 0.98) |
| 7-8 | 0.74 (0.73, 0.74) | 0.75 (0.75, 0.76) | 0.92 (0.90, 0.93) |
| 8-9 | 0.80 (0.79, 0.80) | 0.80 (0.80, 0.81) | 0.91 (0.90, 0.93) |
| ≥10 | 0.87 (0.86, 0.88) | 0.86 (0.86, 0.87) | 0.90 (0.89, 0.92) |
| **NO_2_ category (parts per billion) (ref: 0 – 6)** |  |  |  |
| 6-8 | 0.85 (0.85, 0.86) | 0.87 (0.86, 0.87) | 0.95 (0.95, 0.96) |
| ≥8 | 0.83 (0.83, 0.83) | 0.83 (0.83, 0.84) | 0.94 (0.93, 0.95) |
| **Social determinants of health^h^** |  |  |  |
| **Household density quintile^i^ (ref: 1^st^ quintile)** |  |  |  |
| 2 | 0.90 (0.90, 0.91) | 0.96 (0.95, 0.96) | 1.01 (1.00, 1.02) |
| 3 | 0.86 (0.85, 0.86) | 0.93 (0.92, 0.94) | 1.03 (1.02, 1.04) |
| 4 | 0.78 (0.77, 0.78) | 0.86 (0.86, 0.87) | 1.01 (1.00, 1.02) |
| 5 | 0.64 (0.63, 0.64) | 0.73 (0.73, 0.74) | 0.97 (0.95, 0.98) |
| **Apartment building density category^j^ (ref: 1^st^ category)** |  |  |  |
| 2 | 1.19 (1.18, 1.20) | 1.16 (1.15, 1.16) | 1.04 (1.03, 1.04) |
| 3 | 1.19 (1.18, 1.20) | 1.12 (1.11, 1.13) | 1.01 (1.00, 1.02) |
| **Uncoupled quintile^k^ (ref: 1^st^ quintile)** |  |  |  |
| 2 | 1.00 (0.99, 1.01) | 0.98 (0.97, 0.99) | 1.02 (1.01, 1.03) |
| 3 | 1.05 (1.04, 1.05) | 1.00 (1.00, 1.01) | 1.07 (1.06, 1.08) |
| 4 | 1.17 (1.16, 1.18) | 1.10 (1.09, 1.11) | 1.19 (1.18, 1.21) |
| 5 | 1.36 (1.35, 1.37) | 1.27 (1.26, 1.28) | 1.39 (1.38, 1.41) |
| **Essential work quintile^l^ (ref: 1^st^ quintile)** |  |  |  |
| 2 | 1.03 (1.02, 1.04) | 1.05 (1.04, 1.06) | 1.04 (1.03, 1.04) |
| 3 | 1.13 (1.12, 1.14) | 1.13 (1.13, 1.14) | 1.06 (1.05, 1.07) |
| 4 | 1.13 (1.13, 1.14) | 1.15 (1.14, 1.15) | 1.05 (1.04, 1.06) |
| 5 | 1.15 (1.15, 1.16) | 1.18 (1.17, 1.19) | 1.04 (1.03, 1.06) |
| **Household income quintile^m^ (ref: 1^st^ quintile)** |  |  |  |
| 2 | 0.94 (0.93, 0.94) | 0.93 (0.92, 0.93) | 1.03 (1.02, 1.04) |
| 3 | 0.88 (0.88, 0.89) | 0.89 (0.89, 0.90) | 1.08 (1.07, 1.09) |
| 4 | 0.84 (0.84, 0.85) | 0.86 (0.86, 0.87) | 1.10 (1.09, 1.11) |
| 5 | 0.82 (0.82, 0.83) | 0.84 (0.83, 0.84) | 1.11 (1.09, 1.12) |
| **Limited educational attainment quintile^n^ (ref: 1^st^ quintile)** |  |  |  |
| 2 | 1.05 (1.04, 1.06) | 1.06 (1.05, 1.07) | 1.03 (1.02, 1.04) |
| 3 | 1.07 (1.06, 1.08) | 1.08 (1.07, 1.09) | 1.01 (1.00, 1.02) |
| 4 | 1.14 (1.13, 1.15) | 1.14 (1.13, 1.15) | 1.02 (1.01, 1.03) |
| 5 | 1.15 (1.14, 1.16) | 1.16 (1.16, 1.17) | 1.01 (1.00, 1.02) |
| **Visible minority quintile^o^ (ref: 1^st^ quintile)** |  |  |  |
| 2 | 0.99 (0.98, 1.00) | 0.98 (0.98, 0.99) | 0.99 (0.98, 1.00) |
| 3 | 0.92 (0.91, 0.92) | 0.91 (0.91, 0.92) | 0.95 (0.94, 0.96) |
| 4 | 0.84 (0.83, 0.84) | 0.85 (0.85, 0.86) | 0.91 (0.90, 0.92) |
| 5 | 0.73 (0.72, 0.73) | 0.76 (0.76, 0.77) | 0.86 (0.85, 0.87) |
| **Recent immigration category^p^ (ref: 1^st^ category)** |  |  |  |
| 2 | 0.91 (0.90, 0.91) | 0.92 (0.92, 0.93) | 0.99 (0.99, 1.00) |
| 3 | 0.81 (0.81, 0.82) | 0.84 (0.83, 0.84) | 0.94 (0.93, 0.95) |

^a^ Fully-adjusted model contains all variables listed in this table as covariates.

^b^ “Rural” defined as located outside the commuting zone of a city with greater than 10,000 population.

^c^ Participants were counted if they had a diagnosis in the last 5 years.

^d^ Participants considered “immunocompromised” if they were HIV positive, or had an organ or bone marrow transplant, or another immunodeficient condition.

^e^ This category includes individuals with ischemic stroke or transient ischemic attack in the last 20 years.

^f^ This category includes individuals with a diagnosis in the last 2 years.

^g^ Values of PM_2.5_ > 12µg/m^3^ or NO_2_ > 53ppb yearly have been found to be associated with increased risk of other respiratory illnesses.^80^

^h^ All variables in this category are area-level variables at the level of the Census Dissemination Area.

^i^ 1^st^ quintile represents 0-2.1 persons/dwelling; 2^nd^ quintile: 2.2 – 2.4 persons/dwelling; 3^rd^ quintile: 2.5 – 2.6 persons/dwelling; 4^th^ quintile: 2.7 – 3 persons/dwelling; 5^th^ quintile: 3.1 – 5.7 persons/dwelling.

^j^ 1^st^ category represents 0 – 7.3% of buildings in the area being apartment buildings; 2^nd^ category: 7.4 – 37.7% of buildings; 3^rd^ category: 37.7 – 100% of buildings.

^k^ 1^st^ quintile represents 11.2 – 33.7% individuals uncoupled; 2^nd^ quintile: 33.7 – 38.4% individuals; 3^rd^ quintile: 38.5 – 43.6% individuals; 4^th^ quintile: 43.6 – 51.0% individuals; 5^th^ quintile: 51.0 – 94.6% individuals.

^l^ 1^st^ quintile represents 0 – 32.5% of working individuals in the area self-identifying as working in an essential job; 2^nd^ quintile: 32.5 – 42.3%of individuals; 3^rd^ quintile: 42.3 – 49.8% of individuals; 4^th^ quintile: 50.0 – 57.5% of individuals; 5^th^ quintile: 57.5 – 114.3% of individuals.

^m^ Income quintile has variable cut-off values in each city or census area, to take cost of living into account. A DA being in quintile 1 means it is among the lowest 20% DAs in its city by income.

^n^ 1^st^ quintile represents 0 – 4.1% of individuals 25-64 years old without diploma; 2^nd^ quintile: 4.1 – 7.5% of individuals; 3^rd^ quintile: 7.5 – 11.4% of individuals; 4^th^ quintile: 11.4 – 17.1% of individuals; 5^th^ quintile: 17.1 – 94.3% of individuals.

^o^ 1^st^ quintile represents 0 – 2.2% of individuals in the area self-identifying as a visible minority; 2^nd^ quintile: 2.2 – 7.5% of individuals; 3^rd^ quintile: 7.5 – 18.7% of individuals; 4^th^ quintile: 18.7 – 43.5% of individuals; 5^th^ quintile: 43.5 – 100% of individuals.

^p^ 1^st^ category represents 0 – 2.1% of individuals in DA being recent immigrants; 2^nd^ category: 2.1 – 4.7% of individuals; 3^rd^ category: 4.7 – 41.2% of individuals.
