## Supplemental Table 4 for "The Individual and Social Determinants of COVID-19 Diagnosis in Ontario, Canada: A Population-Wide Study"

**Supplemental Table 4.** Unadjusted, age/sex-adjusted, and fully-adjusted model results describing the odds of COVID-19 diagnosis, according to the pseudo-test-negative design, in Ontario, Canada between March 1 and June 20, 2020.

| **Demographic characteristics** | **Odds of COVID-19 diagnosis, among all individuals tested (pseudo-test-negative design): OR (95% CI)** | | |
| --- | --- | --- | --- |
|  | **Unadjusted** | **Age/sex-adjusted** | **Fully-adjusted^a^** |
| **Sample size** | 758,691 | 758,691 | 758,691 |
| **Age groups (years) (ref: 0 – 4)** |  |  |  |
| 5-19 | 1.66 (1.44, 1.92) | 1.36 (1.33, 1.40) | 1.93 (1.66, 2.24) |
| 20-34 | 1.87 (1.63, 2.14) | 1.72 (1.49, 1.99) | 2.00 (1.75, 2.30) |
| 35-49 | 1.85 (1.61, 2.11) | 1.96 (1.71, 2.25) | 1.98 (1.73, 2.27) |
| 50-64 | 1.78 (1.55, 2.03) | 1.96 (1.71, 2.24) | 2.02 (1.76, 2.32) |
| 65-74 | 1.20 (1.04, 1.38) | 1.88 (1.64, 2.15) | 1.62 (1.40, 1.87) |
| 75-84 | 1.19 (1.03, 1.37) | 1.23 (1.07, 1.42) | 1.64 (1.41, 1.90) |
| ≥85 | 1.36 (1.18, 1.58) | 1.22 (1.05, 1.41) | 1.76 (1.51, 2.06) |
| Male sex | 1.33 (1.30, 1.36) | 1.45 (1.25, 1.67) | 1.26 (1.23, 1.30) |
| Living in rural/small town^b^ | 0.29 (0.27, 0.31) | 0.30 (0.28, 0.32) | 0.81 (0.74, 0.88) |
| **Underlying chronic health conditions** |  |  |  |
| Asthma | 0.80 (0.78, 0.83) | 0.80 (0.78, 0.83) | 0.86 (0.83, 0.89) |
| COPD | 0.60 (0.56, 0.65) | 0.70 (0.65, 0.75) | 0.89 (0.82, 0.96) |
| Hypertension | 0.96 (0.93, 0.98) | 1.19 (1.15, 1.23) | 1.12 (1.08, 1.16) |
| Diabetes | 1.18 (1.14, 1.22) | 1.36 (1.31, 1.41) | 1.26 (1.21, 1.31) |
| Congestive heart failure | 0.79 (0.74, 0.85) | 0.95 (0.89, 1.02) | 0.99 (0.92, 1.07) |
| Dementia or frailty score >15 | 1.02 (0.96, 1.08) | 1.35 (1.26, 1.44) | 1.39 (1.29, 1.49) |
| Cancer^c^ | 0.57 (0.52, 0.63) | 0.65 (0.59, 0.71) | 0.72 (0.66, 0.80) |
| Chronic kidney disease^c^ | 0.87 (0.82, 0.92) | 1.00 (0.93, 1.06) | 0.86 (0.80, 0.93) |
| Immunocompromised^d^ | 0.70 (0.62, 0.78) | 0.72 (0.64, 0.80) | 0.79 (0.70, 0.89) |
| Advanced liver disease | 0.69 (0.60, 0.79) | 0.71 (0.62, 0.82) | 0.79 (0.68, 0.90) |
| Ischemic heart disease | 0.75 (0.70, 0.81) | 0.84 (0.79, 0.91) | 0.88 (0.82, 0.95) |
| Ischemic stroke or transient ischemic attack^e^ | 0.87 (0.79, 0.96) | 1.04 (0.94, 1.15) | 1.05 (0.95, 1.17) |
| Schizophrenia^f^ | 0.86 (0.75, 0.98) | 0.78 (0.68, 0.90) | 0.84 (0.73, 0.97) |
| Substance abuse^f^ | 0.62 (0.57, 0.68) | 0.55 (0.50, 0.60) | 0.72 (0.66, 0.79) |
| **Healthcare use** |  |  |  |
| **Aggregated Diagnostic Group quintile (ref: 0 ADGs)** |  |  |  |
| 2 (1-2 ADGs) | 0.72 (0.68, 0.76) | 0.74 (0.70, 0.79) | 0.78 (0.73, 0.84) |
| 3 (3-4 ADGs) | 0.70 (0.66, 0.74) | 0.75 (0.71, 0.79) | 0.75 (0.70, 0.80) |
| 4 (5-6 ADGs) | 0.69 (0.65, 0.73) | 0.77 (0.73, 0.82) | 0.74 (0.69, 0.79) |
| 5 (7-27 ADGs) | 0.65 (0.62, 0.69) | 0.78 (0.74, 0.83) | 0.75 (0.69, 0.80) |
| **Hospital admissions, past 3 years (ref: 0 admissions)** |  |  |  |
| 1 | 0.75 (0.72, 0.79) | 0.83 (0.79, 0.86) | 0.93 (0.89, 0.97) |
| 2 | 0.67 (0.63, 0.72) | 0.75 (0.70, 0.81) | 0.86 (0.79, 0.93) |
| ≥3 | 0.67 (0.63, 0.72) | 0.74 (0.68, 0.79) | 0.82 (0.76, 0.90) |
| **Outpatient physician visits, past year (ref: 0 – 1 visits)** |  |  |  |
| 2 – 4 | 0.96 (0.93, 1.00) | 1.02 (0.98, 1.06) | 1.02 (0.98, 1.07) |
| 5 – 8 | 1.00 (0.96, 1.04) | 1.11 (1.07, 1.16) | 1.03 (0.98, 1.08) |
| 9 – 14 | 0.97 (0.93, 1.01) | 1.13 (1.08, 1.18) | 0.98 (0.93, 1.04) |
| ≥15 | 0.89 (0.85, 0.93) | 1.07 (1.02, 1.12) | 0.91 (0.85, 0.97) |
| Influenza vaccination (2019-2020 season) | 0.69 (0.67, 0.71) | 0.76 (0.73, 0.78) | 0.81 (0.78, 0.83) |
| **Environmental determinants^g^** |  |  |  |
| **PM_2.5_ category (µg/m^3^) (ref: 2 – 6)** |  |  |  |
| 6-7 | 1.71 (1.60, 1.82) | 1.69 (1.58, 1.81) | 0.91 (0.85, 0.99) |
| 7-8 | 3.59 (3.41, 3.78) | 3.53 (3.35, 3.71) | 1.10 (0.99, 1.21) |
| 8-9 | 4.63 (4.40, 4.86) | 4.50 (4.28, 4.73) | 1.29 (1.16, 1.43) |
| ≥10 | 2.58 (2.41, 2.76) | 2.56 (2.40, 2.74) | 1.45 (1.29, 1.63) |
| **NO_2_ category (parts per billion) (ref: 0 – 6)** |  |  |  |
| 6-8 | 2.06 (1.98, 2.14) | 2.02 (1.95, 2.10) | 1.05 (1.00, 1.11) |
| ≥8 | 3.81 (3.69, 3.93) | 3.70 (3.58, 3.82) | 1.13 (1.06, 1.21) |
| **Social determinants of health^h^** |  |  |  |
| **Household density quintile^i^ (ref: 1^st^ quintile)** |  |  |  |
| 2 | 0.99 (0.94, 1.04) | 0.99 (0.94, 1.03) | 1.21 (1.14, 1.27) |
| 3 | 1.16 (1.10, 1.22) | 1.15 (1.09, 1.21) | 1.39 (1.31, 1.48) |
| 4 | 1.70 (1.63, 1.77) | 1.67 (1.61, 1.74) | 1.70 (1.61, 1.79) |
| 5 | 2.62 (2.51, 2.72) | 2.52 (2.43, 2.63) | 1.94 (1.82, 2.07) |
| **Apartment building density category^j^ (ref: 1^st^ category)** |  |  |  |
| 2 | 0.84 (0.81, 0.88) | 0.86 (0.83, 0.89) | 1.00 (0.95, 1.04) |
| 3 | 1.41 (1.37, 1.45) | 1.42 (1.38, 1.46) | 1.15 (1.09, 1.21) |
| **Uncoupled quintile^k^ (ref: 1^st^ quintile)** |  |  |  |
| 2 | 1.29 (1.23, 1.35) | 1.29 (1.23, 1.35) | 0.96 (0.91, 1.01) |
| 3 | 1.54 (1.47, 1.61) | 1.54 (1.47, 1.61) | 0.95 (0.90, 1.00) |
| 4 | 1.63 (1.56, 1.70) | 1.64 (1.57, 1.71) | 0.96 (0.91, 1.02) |
| 5 | 1.70 (1.64, 1.78) | 1.72 (1.65, 1.79) | 1.07 (1.01, 1.15) |
| **Essential work quintile^l^ (ref: 1^st^ quintile)** |  |  |  |
| 2 | 1.21 (1.16, 1.26) | 1.21 (1.16, 1.26) | 1.25 (1.19, 1.32) |
| 3 | 1.16 (1.11, 1.21) | 1.17 (1.12, 1.22) | 1.28 (1.21, 1.35) |
| 4 | 1.34 (1.29, 1.40) | 1.36 (1.30, 1.41) | 1.37 (1.29, 1.45) |
| 5 | 1.61 (1.54, 1.67) | 1.62 (1.55, 1.69) | 1.42 (1.32, 1.51) |
| **Household income quintile^m^ (ref: 1^st^ quintile)** |  |  |  |
| 2 | 0.79 (0.77, 0.82) | 0.80 (0.77, 0.83) | 0.96 (0.91, 1.00) |
| 3 | 0.78 (0.75, 0.81) | 0.78 (0.75, 0.81) | 1.02 (0.97, 1.08) |
| 4 | 0.64 (0.61, 0.67) | 0.64 (0.61, 0.66) | 0.97 (0.90, 1.03) |
| 5 | 0.55 (0.53, 0.58) | 0.55 (0.53, 0.57) | 0.97 (0.90, 1.04) |
| **Limited educational attainment quintile^n^ (ref: 1^st^ quintile)** |  |  |  |
| 2 | 1.07 (1.03, 1.12) | 1.08 (1.04, 1.13) | 1.07 (1.02, 1.12) |
| 3 | 1.23 (1.18, 1.29) | 1.24 (1.19, 1.29) | 1.15 (1.09, 1.21) |
| 4 | 1.33 (1.27, 1.38) | 1.34 (1.29, 1.40) | 1.23 (1.17, 1.30) |
| 5 | 1.72 (1.65, 1.79) | 1.72 (1.65, 1.79) | 1.37 (1.29, 1.46) |
| **Visible minority quintile^o^ (ref: 1^st^ quintile)** |  |  |  |
| 2 | 1.24 (1.16, 1.32) | 1.23 (1.16, 1.32) | 0.96 (0.89, 1.02) |
| 3 | 1.77 (1.67, 1.88) | 1.75 (1.65, 1.86) | 0.97 (0.91, 1.05) |
| 4 | 2.72 (2.57, 2.87) | 2.66 (2.52, 2.81) | 1.07 (0.99, 1.15) |
| 5 | 5.52 (5.24, 5.81) | 5.34 (5.07, 5.62) | 1.27 (1.17, 1.38) |
| **Recent immigration category^p^ (ref: 1^st^ category)** |  |  |  |
| 2 | 1.79 (1.73, 1.85) | 1.76 (1.70, 1.82) | 1.04 (1.00, 1.09) |
| 3 | 3.06 (2.97, 3.15) | 2.97 (2.89, 3.06) | 1.16 (1.11, 1.22) |
