## Supplemental Table 5 for "The Individual and Social Determinants of COVID-19 Diagnosis in Ontario, Canada: A Population-Wide Study"

**Supplemental Table 5.** Unadjusted, age/sex-adjusted, and fully-adjusted model results describing the odds of COVID-19 diagnosis, according to the true test-negative design, in Ontario, Canada between March 1 and June 20, 2020.

| **Demographic characteristics** | **Odds of COVID-19 diagnosis among symptomatic tested individuals only (true test-negative design): OR (95% CI)** | | |
| --- | --- | --- | --- |
|  | **Unadjusted** | **Age/sex-adjusted** | **Fully-adjusted^a^** |
| **Sample size** | 89,408 | 89,408 | 89,408 |
| **Age groups (years) (ref: 0 – 4)** |  |  |  |
| 5-19 | 2.98 (1.79, 4.96) | 1.43 (1.34, 1.52) | 2.95 (1.76, 4.93) |
| 20-34 | 4.25 (2.63, 6.89) | 3.06 (1.84, 5.10) | 4.03 (2.47, 6.57) |
| 35-49 | 4.36 (2.69, 7.06) | 4.50 (2.78, 7.30) | 4.21 (2.58, 6.85) |
| 50-64 | 5.20 (3.21, 8.42) | 4.65 (2.87, 7.54) | 5.48 (3.36, 8.93) |
| 65-74 | 4.21 (2.59, 6.87) | 5.53 (3.42, 8.96) | 5.09 (3.09, 8.37) |
| 75-84 | 4.64 (2.83, 7.60) | 4.36 (2.68, 7.11) | 5.42 (3.26, 9.02) |
| ≥85 | 6.73 (4.12, 10.99) | 4.80 (2.93, 7.87) | 6.01 (3.61, 10.03) |
| Male sex | 1.39 (1.31, 1.48) | 7.24 (4.43, 11.83) | 1.35 (1.26, 1.44) |
| Living in rural/small town^b^ | 0.41 (0.36, 0.46) | 0.41 (0.37, 0.46) | 0.76 (0.65, 0.88) |
| **Underlying chronic health conditions** |  |  |  |
| Asthma | 0.75 (0.69, 0.82) | 0.77 (0.71, 0.84) | 0.85 (0.78, 0.93) |
| COPD | 0.76 (0.65, 0.89) | 0.64 (0.55, 0.76) | 0.83 (0.70, 0.98) |
| Hypertension | 1.20 (1.12, 1.28) | 1.04 (0.95, 1.13) | 1.10 (1.00, 1.20) |
| Diabetes | 1.27 (1.17, 1.38) | 1.14 (1.05, 1.25) | 1.18 (1.07, 1.30) |
| Congestive heart failure | 1.06 (0.91, 1.23) | 0.83 (0.71, 0.98) | 0.92 (0.77, 1.10) |
| Dementia or frailty score >15 | 1.90 (1.68, 2.15) | 1.76 (1.52, 2.04) | 1.70 (1.44, 1.99) |
| Cancer^c^ | 0.80 (0.65, 0.99) | 0.72 (0.58, 0.89) | 0.88 (0.70, 1.10) |
| Chronic kidney disease^c^ | 1.18 (1.02, 1.37) | 1.01 (0.87, 1.18) | 1.04 (0.88, 1.23) |
| Immunocompromised^d^ | 0.80 (0.61, 1.04) | 0.76 (0.58, 0.99) | 0.84 (0.64, 1.11) |
| Advanced liver disease | 0.70 (0.50, 0.99) | 0.63 (0.45, 0.89) | 0.76 (0.53, 1.07) |
| Ischemic heart disease | 0.94 (0.80, 1.09) | 0.74 (0.63, 0.87) | 0.81 (0.68, 0.97) |
| Ischemic stroke or transient ischemic attack^e^ | 1.06 (0.84, 1.34) | 0.88 (0.69, 1.11) | 0.94 (0.73, 1.20) |
| Schizophrenia^f^ | 1.21 (0.89, 1.65) | 1.12 (0.82, 1.53) | 1.27 (0.92, 1.75) |
| Substance abuse^f^ | 0.41 (0.31, 0.53) | 0.39 (0.30, 0.50) | 0.52 (0.40, 0.68) |
| **Healthcare use** |  |  |  |
| **Aggregated Diagnostic Group quintile (ref: 0 ADGs)** |  |  |  |
| 2 (1-2 ADGs) | 0.63 (0.54, 0.74) | 0.65 (0.56, 0.76) | 0.68 (0.58, 0.80) |
| 3 (3-4 ADGs) | 0.55 (0.48, 0.64) | 0.57 (0.49, 0.66) | 0.57 (0.48, 0.68) |
| 4 (5-6 ADGs) | 0.56 (0.49, 0.65) | 0.58 (0.50, 0.67) | 0.56 (0.47, 0.67) |
| 5 (7-27 ADGs) | 0.54 (0.47, 0.62) | 0.53 (0.46, 0.61) | 0.52 (0.43, 0.63) |
| **Hospital admissions, past 3 years (ref: 0 admissions)** |  |  |  |
| 1 | 0.74 (0.67, 0.81) | 0.74 (0.67, 0.83) | 0.88 (0.79, 0.98) |
| 2 | 0.69 (0.58, 0.83) | 0.64 (0.53, 0.77) | 0.80 (0.66, 0.97) |
| ≥3 | 0.92 (0.79, 1.08) | 0.81 (0.69, 0.95) | 1.03 (0.85, 1.25) |
| **Outpatient physician visits, past year (ref: 0 – 1 visits)** |  |  |  |
| 2 – 4 | 0.96 (0.87, 1.05) | 0.97 (0.88, 1.06) | 1.07 (0.95, 1.19) |
| 5 – 8 | 0.93 (0.85, 1.02) | 0.93 (0.84, 1.03) | 1.01 (0.89, 1.15) |
| 9 – 14 | 0.98 (0.89, 1.09) | 0.97 (0.87, 1.07) | 1.04 (0.90, 1.20) |
| ≥15 | 0.93 (0.83, 1.04) | 0.87 (0.78, 0.98) | 0.88 (0.75, 1.03) |
| Influenza vaccination (2019-2020 season) | 0.89 (0.83, 0.95) | 0.83 (0.77, 0.90) | 0.86 (0.79, 0.93) |
| **Environmental determinants^g^** |  |  |  |
| **PM_2.5_ category (µg/m^3^) (ref: 2 – 6)** |  |  |  |
| 6-7 | 1.45 (1.26, 1.67) | 1.45 (1.26, 1.67) | 0.81 (0.69, 0.95) |
| 7-8 | 3.02 (2.73, 3.34) | 3.00 (2.71, 3.32) | 1.00 (0.81, 1.23) |
| 8-9 | 3.47 (3.13, 3.83) | 3.46 (3.13, 3.83) | 1.17 (0.94, 1.45) |
| ≥10 | 4.49 (3.97, 5.08) | 4.45 (3.93, 5.03) | 1.52 (1.20, 1.93) |
| **NO_2_ category (parts per billion) (ref: 0 – 6)** |  |  |  |
| 6-8 | 2.00 (1.85, 2.17) | 1.99 (1.84, 2.16) | 1.04 (0.94, 1.16) |
| ≥8 | 2.45 (2.28, 2.64) | 2.42 (2.25, 2.61) | 1.09 (0.94, 1.26) |
| **Social determinants of health^h^** |  |  |  |
| **Household density quintile^i^ (ref: 1^st^ quintile)** |  |  |  |
| 2 | 1.02 (0.91, 1.14) | 1.04 (0.94, 1.16) | 1.23 (1.08, 1.38) |
| 3 | 1.21 (1.08, 1.36) | 1.25 (1.12, 1.41) | 1.33 (1.16, 1.53) |
| 4 | 1.83 (1.66, 2.02) | 1.90 (1.72, 2.10) | 1.74 (1.52, 2.00) |
| 5 | 2.13 (1.93, 2.35) | 2.20 (1.99, 2.43) | 1.66 (1.41, 1.96) |
| **Apartment building density category^j^ (ref: 1^st^ category)** |  |  |  |
| 2 | 0.80 (0.73, 0.87) | 0.79 (0.73, 0.86) | 1.00 (0.90, 1.11) |
| 3 | 1.12 (1.04, 1.21) | 1.09 (1.01, 1.18) | 1.09 (0.95, 1.24) |
| **Uncoupled quintile^k^ (ref: 1^st^ quintile)** |  |  |  |
| 2 | 1.25 (1.13, 1.39) | 1.24 (1.12, 1.37) | 1.08 (0.97, 1.20) |
| 3 | 1.17 (1.06, 1.30) | 1.15 (1.04, 1.28) | 1.00 (0.88, 1.12) |
| 4 | 1.10 (0.99, 1.22) | 1.09 (0.98, 1.21) | 0.95 (0.82, 1.09) |
| 5 | 1.28 (1.16, 1.40) | 1.24 (1.13, 1.37) | 1.05 (0.90, 1.24) |
| **Essential work quintile^l^ (ref: 1^st^ quintile)** |  |  |  |
| 2 | 1.10 (1.00, 1.22) | 1.11 (1.00, 1.23) | 1.25 (1.11, 1.40) |
| 3 | 0.94 (0.85, 1.05) | 0.96 (0.86, 1.06) | 1.20 (1.05, 1.38) |
| 4 | 0.95 (0.85, 1.05) | 0.97 (0.87, 1.07) | 1.19 (1.03, 1.38) |
| 5 | 1.08 (0.98, 1.20) | 1.11 (1.00, 1.23) | 1.20 (1.03, 1.42) |
| **Household income quintile^m^ (ref: 1^st^ quintile)** |  |  |  |
| 2 | 0.89 (0.80, 0.98) | 0.89 (0.80, 0.98) | 0.95 (0.85, 1.07) |
| 3 | 0.90 (0.82, 1.00) | 0.90 (0.82, 1.00) | 0.96 (0.83, 1.11) |
| 4 | 0.85 (0.77, 0.94) | 0.85 (0.77, 0.94) | 0.93 (0.79, 1.09) |
| 5 | 1.08 (0.98, 1.19) | 1.08 (0.98, 1.18) | 1.22 (1.02, 1.46) |
| **Limited educational attainment quintile^n^ (ref: 1^st^ quintile)** |  |  |  |
| 2 | 1.06 (0.95, 1.17) | 1.05 (0.95, 1.16) | 1.08 (0.97, 1.21) |
| 3 | 0.92 (0.83, 1.02) | 0.93 (0.84, 1.04) | 1.15 (1.02, 1.30) |
| 4 | 1.01 (0.91, 1.12) | 1.03 (0.93, 1.14) | 1.40 (1.22, 1.59) |
| 5 | 1.10 (0.99, 1.22) | 1.11 (1.00, 1.23) | 1.44 (1.24, 1.67) |
| **Visible minority quintile^o^ (ref: 1^st^ quintile)** |  |  |  |
| 2 | 1.34 (1.19, 1.50) | 1.34 (1.19, 1.50) | 1.03 (0.91, 1.17) |
| 3 | 1.90 (1.70, 2.13) | 1.89 (1.70, 2.12) | 1.06 (0.92, 1.21) |
| 4 | 2.57 (2.30, 2.86) | 2.57 (2.30, 2.86) | 1.12 (0.96, 1.31) |
| 5 | 3.24 (2.92, 3.60) | 3.26 (2.94, 3.62) | 1.17 (0.97, 1.42) |
| **Recent immigration category^p^ (ref: 1^st^ category)** |  |  |  |
| 2 | 1.57 (1.45, 1.71) | 1.58 (1.45, 1.71) | 1.13 (1.02, 1.24) |
| 3 | 2.16 (2.00, 2.32) | 2.17 (2.01, 2.34) | 1.31 (1.17, 1.47) |
