## Supplemental Table 6 for "The Individual and Social Determinants of COVID-19 Diagnosis in Ontario, Canada: A Population-Wide Study"

**Supplemental Table 6.** Unadjusted, age/sex-adjusted, and fully-adjusted model results describing the odds of COVID-19 diagnosis, according to the case-control design, in Ontario, Canada between March 1 and June 20, 2020.

| **Demographic characteristics** | **Odds of COVID-19 diagnosis, comparing all test-positive individuals to all test-negatives and all untested individuals (case-control design): OR (95% CI)** | | |
| --- | --- | --- | --- |
|  | **Unadjusted** | **Age/sex-adjusted** | **Fully-adjusted^a^** |
| **Sample size** | 14,695,579 | 14,695,579 | 14,695,579 |
| **Age groups (years) (ref: 0 – 4)** |  |  |  |
| 5-19 | 1.48 (1.28, 1.71) | 1.48 (1.28, 1.71) | 1.72 (1.49, 1.99) |
| 20-34 | 6.38 (5.58, 7.30) | 6.37 (5.57, 7.29) | 7.03 (6.13, 8.05) |
| 35-49 | 6.52 (5.70, 7.46) | 6.50 (5.68, 7.44) | 6.74 (5.88, 7.73) |
| 50-64 | 7.19 (6.28, 8.22) | 7.17 (6.27, 8.21) | 7.01 (6.11, 8.03) |
| 65-74 | 4.40 (3.83, 5.05) | 4.38 (3.81, 5.04) | 3.99 (3.46, 4.60) |
| 75-84 | 5.25 (4.54, 6.05) | 5.22 (4.52, 6.02) | 3.94 (3.39, 4.57) |
| ≥85 | 12.44 (10.77, 14.37) | 12.31 (10.65, 14.21) | 7.26 (6.23, 8.46) |
| Male sex | 0.89 (0.87, 0.92) | 0.91 (0.89, 0.94) | 1.00 (0.98, 1.03) |
| Living in rural/small town^b^ | 0.33 (0.31, 0.35) | 0.33 (0.31, 0.35) | 0.76 (0.70, 0.82) |
| **Underlying chronic health conditions** |  |  |  |
| Asthma | 1.02 (0.99, 1.06) | 1.02 (0.99, 1.06) | 0.92 (0.89, 0.96) |
| COPD | 1.48 (1.37, 1.59) | 1.24 (1.15, 1.34) | 1.04 (0.96, 1.13) |
| Hypertension | 1.61 (1.56, 1.65) | 1.47 (1.42, 1.52) | 1.13 (1.09, 1.17) |
| Diabetes | 1.83 (1.77, 1.89) | 1.64 (1.59, 1.70) | 1.19 (1.14, 1.23) |
| Congestive heart failure | 2.24 (2.10, 2.39) | 1.82 (1.70, 1.95) | 1.25 (1.16, 1.35) |
| Dementia or frailty score >15 | 4.27 (4.02, 4.54) | 3.66 (3.42, 3.93) | 2.60 (2.42, 2.80) |
| Cancer^c^ | 1.19 (1.09, 1.31) | 1.03 (0.94, 1.13) | 0.86 (0.78, 0.94) |
| Chronic kidney disease^c^ | 2.30 (2.16, 2.44) | 1.96 (1.84, 2.09) | 1.16 (1.08, 1.24) |
| Immunocompromised^d^ | 1.54 (1.38, 1.72) | 1.43 (1.28, 1.60) | 1.02 (0.91, 1.14) |
| Advanced liver disease | 1.42 (1.24, 1.62) | 1.23 (1.08, 1.41) | 0.87 (0.76, 1.00) |
| Ischemic heart disease | 1.45 (1.35, 1.55) | 1.26 (1.17, 1.35) | 0.90 (0.84, 0.97) |
| Ischemic stroke or transient ischemic attack^e^ | 2.13 (1.93, 2.35) | 1.70 (1.54, 1.88) | 1.18 (1.06, 1.31) |
| Schizophrenia^f^ | 1.80 (1.57, 2.05) | 1.53 (1.34, 1.75) | 0.99 (0.87, 1.13) |
| Substance abuse^f^ | 1.18 (1.08, 1.29) | 1.03 (0.94, 1.12) | 0.86 (0.79, 0.95) |
| **Healthcare use** |  |  |  |
| **Aggregated Diagnostic Group quintile (ref: 0 ADGs)** |  |  |  |
| 2 (1-2 ADGs) | 1.17 (1.10, 1.24) | 1.25 (1.18, 1.33) | 1.24 (1.17, 1.32) |
| 3 (3-4 ADGs) | 1.52 (1.44, 1.61) | 1.63 (1.54, 1.73) | 1.46 (1.37, 1.56) |
| 4 (5-6 ADGs) | 1.93 (1.83, 2.05) | 2.03 (1.91, 2.15) | 1.69 (1.57, 1.81) |
| 5 (7-27 ADGs) | 2.92 (2.77, 3.08) | 2.91 (2.76, 3.08) | 2.12 (1.97, 2.28) |
| **Hospital admissions, past 3 years (ref: 0 admissions)** |  |  |  |
| 1 | 1.06 (1.01, 1.10) | 1.14 (1.09, 1.18) | 0.92 (0.88, 0.96) |
| 2 | 1.46 (1.36, 1.57) | 1.40 (1.31, 1.51) | 1.01 (0.94, 1.09) |
| ≥3 | 2.81 (2.62, 3.02) | 2.54 (2.37, 2.73) | 1.38 (1.27, 1.50) |
| **Outpatient physician visits, past year (ref: 0 – 1 visits)** |  |  |  |
| 2 – 4 | 1.46 (1.41, 1.52) | 1.48 (1.43, 1.54) | 1.12 (1.08, 1.18) |
| 5 – 8 | 1.94 (1.87, 2.01) | 1.92 (1.85, 2.00) | 1.17 (1.11, 1.23) |
| 9 – 14 | 2.37 (2.27, 2.47) | 2.29 (2.20, 2.39) | 1.17 (1.10, 1.24) |
| ≥15 | 3.04 (2.91, 3.18) | 2.80 (2.67, 2.93) | 1.22 (1.14, 1.30) |
| Influenza vaccination (2019-2020 season) | 1.02 (0.99, 1.05) | 0.95 (0.92, 0.98) | 0.87 (0.85, 0.90) |
| **Environmental determinants^g^** |  |  |  |
| **PM_2.5_ category (µg/m^3^) (ref: 2 – 6)** |  |  |  |
| 6-7 | 1.55 (1.45, 1.65) | 1.54 (1.44, 1.64) | 0.92 (0.85, 0.99) |
| 7-8 | 2.62 (2.49, 2.76) | 2.65 (2.52, 2.79) | 1.00 (0.91, 1.10) |
| 8-9 | 3.59 (3.42, 3.77) | 3.58 (3.41, 3.76) | 1.19 (1.08, 1.32) |
| ≥10 | 2.22 (2.08, 2.37) | 2.21 (2.07, 2.36) | 1.31 (1.16, 1.47) |
| **NO_2_ category (parts per billion) (ref: 0 – 6)** |  |  |  |
| 6-8 | 1.74 (1.68, 1.81) | 1.75 (1.69, 1.82) | 1.00 (0.96, 1.06) |
| ≥8 | 3.06 (2.96, 3.16) | 3.05 (2.95, 3.14) | 1.05 (0.98, 1.12) |
| **Social determinants of health^h^** |  |  |  |
| **Household density quintile^i^ (ref: 1^st^ quintile)** |  |  |  |
| 2 | 0.90 (0.85, 0.94) | 0.94 (0.90, 0.99) | 1.19 (1.13, 1.26) |
| 3 | 1.00 (0.95, 1.05) | 1.07 (1.02, 1.13) | 1.42 (1.34, 1.50) |
| 4 | 1.33 (1.27, 1.38) | 1.44 (1.38, 1.50) | 1.70 (1.61, 1.80) |
| 5 | 1.65 (1.58, 1.71) | 1.85 (1.78, 1.92) | 1.86 (1.75, 1.98) |
| **Apartment building density category^j^ (ref: 1^st^ category)** |  |  |  |
| 2 | 1.00 (0.96, 1.04) | 0.98 (0.94, 1.02) | 1.02 (0.98, 1.06) |
| 3 | 1.64 (1.60, 1.69) | 1.56 (1.52, 1.60) | 1.18 (1.12, 1.24) |
| **Uncoupled quintile^k^ (ref: 1^st^ quintile)** |  |  |  |
| 2 | 1.28 (1.23, 1.34) | 1.26 (1.20, 1.32) | 0.97 (0.92, 1.02) |
| 3 | 1.59 (1.52, 1.66) | 1.53 (1.47, 1.60) | 0.99 (0.94, 1.04) |
| 4 | 1.87 (1.79, 1.94) | 1.77 (1.70, 1.85) | 1.11 (1.05, 1.17) |
| 5 | 2.24 (2.15, 2.34) | 2.11 (2.02, 2.19) | 1.41 (1.32, 1.51) |
| **Essential work quintile^l^ (ref: 1^st^ quintile)** |  |  |  |
| 2 | 1.24 (1.19, 1.29) | 1.26 (1.21, 1.31) | 1.30 (1.24, 1.37) |
| 3 | 1.29 (1.24, 1.35) | 1.30 (1.25, 1.36) | 1.37 (1.30, 1.45) |
| 4 | 1.50 (1.44, 1.56) | 1.52 (1.46, 1.58) | 1.51 (1.42, 1.60) |
| 5 | 1.82 (1.74, 1.89) | 1.85 (1.77, 1.92) | 1.58 (1.48, 1.69) |
| **Household income quintile^m^ (ref: 1^st^ quintile)** |  |  |  |
| 2 | 0.75 (0.73, 0.78) | 0.75 (0.72, 0.77) | 1.00 (0.96, 1.05) |
| 3 | 0.70 (0.68, 0.73) | 0.71 (0.68, 0.73) | 1.12 (1.06, 1.18) |
| 4 | 0.55 (0.53, 0.57) | 0.56 (0.54, 0.58) | 1.06 (1.00, 1.13) |
| 5 | 0.47 (0.45, 0.49) | 0.48 (0.46, 0.45) | 1.07 (0.99, 1.15) |
| **Limited educational attainment quintile^n^ (ref: 1^st^ quintile)** |  |  |  |
| 2 | 1.12 (1.08, 1.17) | 1.13 (1.09, 1.18) | 1.09 (1.04, 1.15) |
| 3 | 1.31 (1.25, 1.36) | 1.32 (1.27, 1.38) | 1.15 (1.09, 1.21) |
| 4 | 1.49 (1.43, 1.55) | 1.49 (1.44, 1.56) | 1.21 (1.15, 1.28) |
| 5 | 1.92 (1.84, 2.00) | 1.95 (1.87, 2.03) | 1.33 (1.26, 1.41) |
| **Visible minority quintile^o^ (ref: 1^st^ quintile)** |  |  |  |
| 2 | 1.22 (1.15, 1.30) | 1.22 (1.14, 1.30) | 0.95 (0.89, 1.02) |
| 3 | 1.62 (1.53, 1.72) | 1.61 (1.52, 1.71) | 0.93 (0.87, 1.00) |
| 4 | 2.25 (2.13, 2.38) | 2.27 (2.15, 2.40) | 0.98 (0.91, 1.06) |
| 5 | 3.87 (3.68, 4.07) | 4.00 (3.80, 4.20) | 1.09 (1.00, 1.19) |
| **Recent immigration category^p^ (ref: 1^st^ category)** |  |  |  |
| 2 | 1.60 (1.55, 1.66) | 1.62 (1.57, 1.68) | 1.04 (1.00, 1.08) |
| 3 | 2.41 (2.34, 2.48) | 2.46 (2.39, 2.53) | 1.10 (1.05, 1.15) |
