## Supplemental Table 7 for "The Individual and Social Determinants of COVID-19 Diagnosis in Ontario, Canada: A Population-Wide Study"

**Supplemental Table 7.** Tolerance and variance inflation factors for the fully-adjusted case-control logistic regression model of the odds of COVID-19 diagnosis.

| **Demographic characteristics** | **Tolerance** | **Variance inflation factor** |
| --- | --- | --- |
| Age | 0.60 | 1.67 |
| Male sex | 0.96 | 1.04 |
| Living in rural/small town | 0.71 | 1.41 |
| **Underlying chronic health conditions** |  |  |
| Asthma | 0.97 | 1.03 |
| COPD | 0.92 | 1.09 |
| Hypertension | 0.61 | 1.65 |
| Diabetes | 0.81 | 1.24 |
| Congestive heart failure | 0.84 | 1.19 |
| Dementia or frailty score >15 | 0.91 | 1.10 |
| Cancer | 0.94 | 1.06 |
| Chronic kidney disease | 0.88 | 1.13 |
| Immunocompromised | 0.97 | 1.03 |
| Advanced liver disease | 0.98 | 1.02 |
| Ischemic heart disease | 0.85 | 1.17 |
| Ischemic stroke or transient ischemic attack | 0.95 | 1.05 |
| Schizophrenia | 0.98 | 1.02 |
| Substance abuse | 0.96 | 1.04 |
| **Healthcare use** |  |  |
| Number of Adjusted Diagnostic Groups | 0.45 | 2.20 |
| Hospital admissions, past 3 years | 0.76 | 1.32 |
| Outpatient physician visits, past year | 0.50 | 2.02 |
| Influenza vaccination (2019-2020 season) | 0.86 | 1.16 |
| **Environmental determinants** |  |  |
| PM_2.5_ category | 0.51 | 1.97 |
| NO_2_ category | 0.27 | 3.77 |
| **Social determinants of health** |  |  |
| Household density quintile | 0.22 | 4.55 |
| Apartment building density category | 0.25 | 3.96 |
| Uncoupled quintile | 0.31 | 3.24 |
| Essential work quintile | 0.40 | 2.48 |
| Household income quintile | 0.29 | 3.41 |
| Limited educational attainment quintile | 0.47 | 2.12 |
| Visible minority quintile | 0.21 | 4.76 |
| Recent immigration category | 0.40 | 2.51 |
