## Supplemental Figure 1 for "The Individual and Social Determinants of COVID-19 Diagnosis in Ontario, Canada: A Population-Wide Study"

**Supplemental Figure 1.** Potential for selection bias, via collider mechanism, in the pseudo-test-negative (A), true test-negative (B), and case-control (C) study designs.[52]

A)

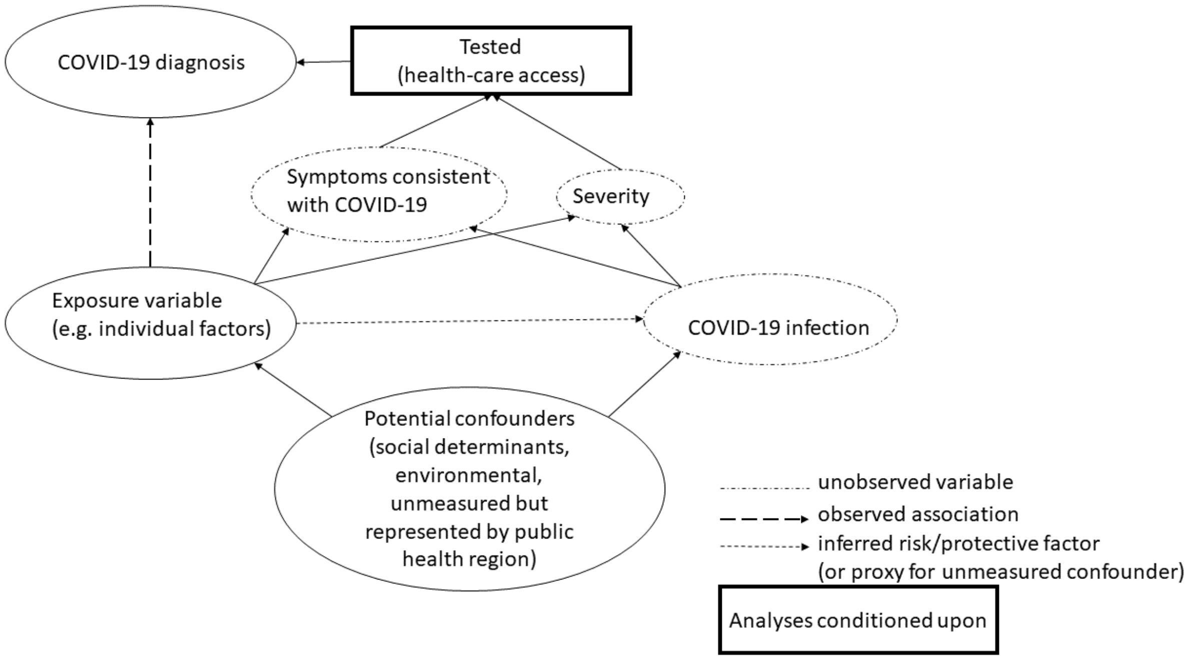


B)

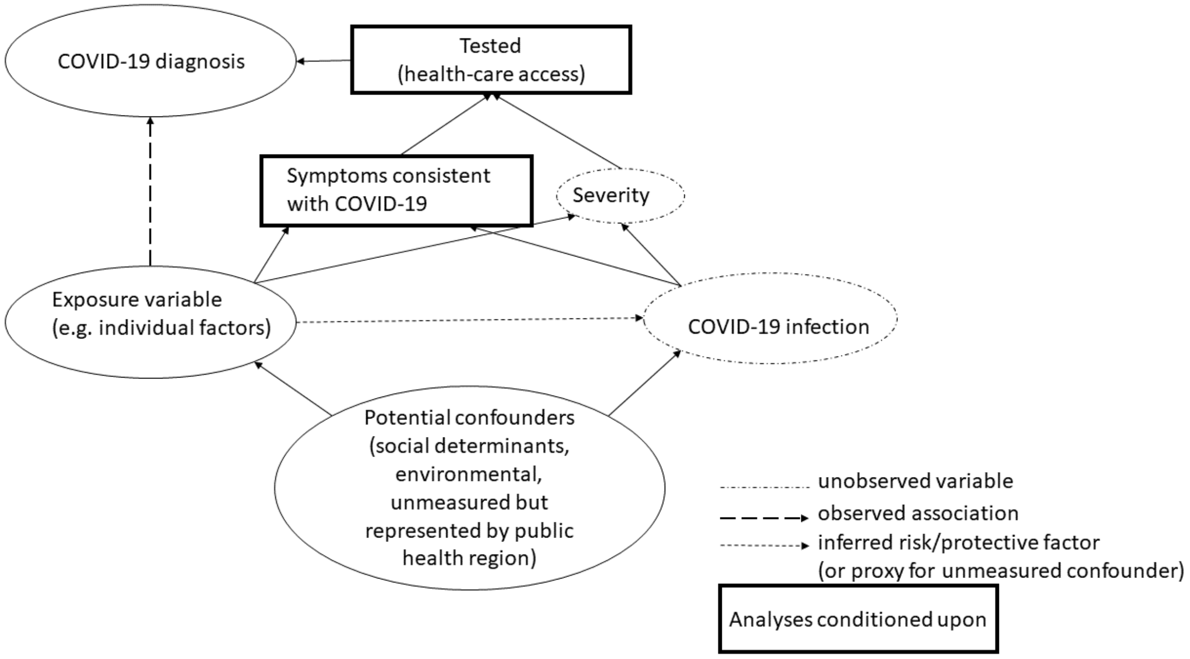

C)

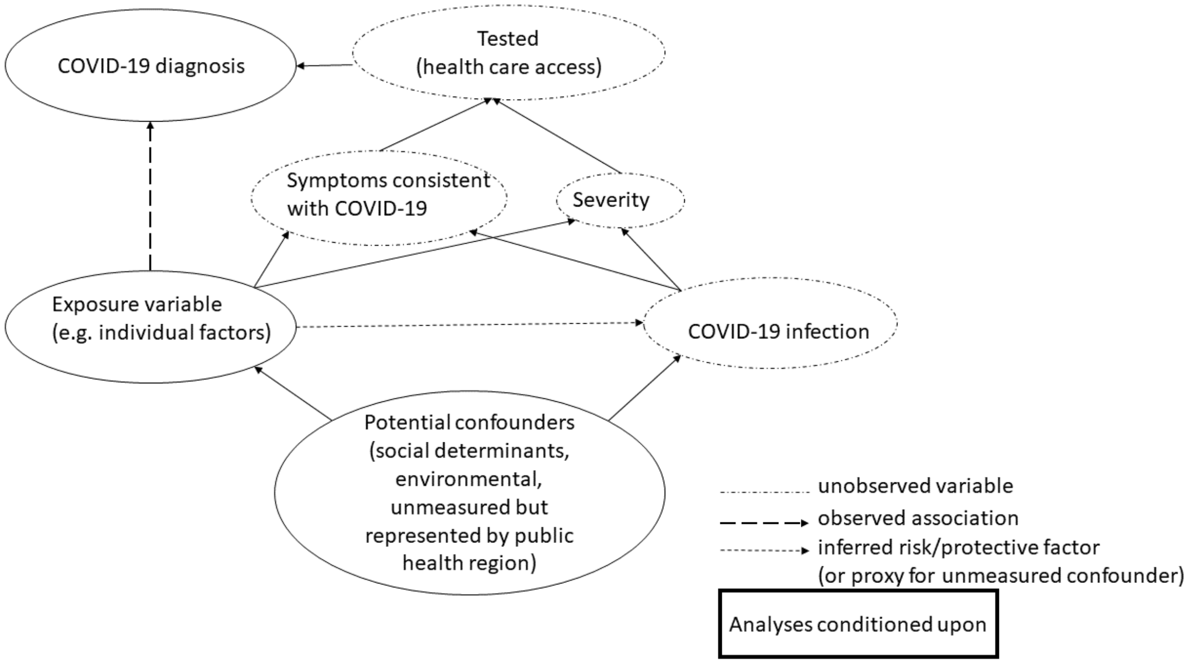


Representative directed acyclic graphs are shown for the exposure category of individual determinants (age, sex or gender, underlying health conditions, prior health care use), to demonstrate the potential mechanisms by which collider bias, a type of selection bias, may influence the relationship between the observed exposure and observed outcome (COVID-19 diagnosis). Panel A) ("pseudo-test-negative design", comparing individuals who tested positive vs. negative) conditions on individuals who are tested for SARS-CoV-2. It has been proposed that when conditioning on testing, individual determinants that may be associated with symptoms consistent with COVID-19 (for example, underlying respiratory or cardiac conditions), severity (for example, age), or different criteria for testing (for example, symptom-based testing vs. asymptomatic/routine screening), may lead to erroneous inference of the direction and magnitude of determinants.[23] Panel B) ("true test-negative design", comparing symptomatic individuals who tested positive vs. negative) is similar to Panel A, but analyses condition on testing in the presence of symptoms consistent with COVID-19. Panel C) ("case-control design", comparing individuals who test positive vs. all others, including those who test negative or have not yet been tested) does not condition on testing. The case-control design implicitly assumes that individuals who were not tested were similar to those who tested negative. Testing assumes access to and/or seeking out of health care. Analyses were conducted with independent directed acyclic graphs for each category (environmental, social determinants) and which were the same as the graphs shown here.
